# How does the macroenvironment influence brain and behaviour – a review of current status and future perspectives

**DOI:** 10.1101/2023.10.09.23296785

**Authors:** Elli Polemiti, Soeren Hese, Kerstin Schepanski, Jiacan Yuan, Gunter Schumann, environMENTAL consortium

**Affiliations:** Centre of Population Neuroscience and Stratified Medicine (PONS), Department of Psychiatry and Clinical Neuroscience, Charité, Universitätsmedizin Berlin, Germany; Institute of Geography, Friedrich Schiller University Jena, Germany; Institute of Meteorology, Free University Berlin, Germany; Department of Atmospheric and Oceanic Sciences & Institute of Atmospheric Sciences & CMA-FDU Joint Laboratory of Marine Meteorology & IRDR-ICOE on Risk Interconnectivity and Governance on Weather/Climate Extremes Impact and Public Health, Fudan University, Shanghai, China; Centre for Population Neuroscience and Precision Medicine (PONS), Institute for Science and Technology of Brain-inspired Intelligence (ISTBI), Fudan University, Shanghai, China

**Author notes:** **Corresponding author: Elli Polemiti, PhD,** Centre for Population Neuroscience and Stratified Medicine (PONS), Department of Psychiatry and Neuroscience, Charite Universitaetsmedizin Berlin (CCM), Chariteplatz 1, 10117 Berlin, Germany, and **Professor Gunter Schumann, MD, PhD**, Centre for Population Nuroscience and Stratified Medicine (PONS), Institute for Science and Technology of Brain-inspired Intelligence (ISTBI), Fudan University, Shanghai, China and Department of Psychiatry and Neuroscience, Charite Universitaetsmedizin Berlin (CCM), Chariteplatz 1, 10117 Berlin, Germany, and.

## Abstract

The environment influences mental health, both detrimentally and beneficially. Current research has emphasized the individual psychosocial ‘microenvironment’. Less attention has been paid to ‘macro-environmental’ challenges including climate change, pollution, urbanicity and socioeconomic disparity. With the advent of large-scale big-data cohorts and an increasingly dense mapping of macroenvironmental parameters, we are now in a position to characterise the relation between macroenvironment, brain, and behaviour across different geographic and cultural locations globally. This review synthesises findings from recent epidemiological and neuroimaging studies, aiming to provide a comprehensive overview of the existing evidence between the macroenvironment and the structure and functions of the brain, with a particular emphasis on its implications for mental illness. We discuss putative underlying mechanisms and address the most common exposures of the macroenvironment. Finally, we identify critical areas for future research to enhance our understanding of the aetiology of mental illness and to inform effective interventions for healthier environments and mental health promotion.

## Introduction

The environment refers to the broader ecological context in which an individual exists, interacts, and adapts (1), and may have direct and indirect effects on mental health (2). It can be broadly divided into the “macroenvironment”, encompassing environmental characteristics at the neighbourhood or larger level, and the “microenvironment”, which relates to the individual psychosocial level (3). The macroenvironment includes factors such as urbanisation, climate patterns, geological features, and ecosystem interactions, as well as socioeconomic disparity – all of which are undergoing rapid and dynamic changes. Urbanisation continues at unprecedented rates, with more than 50% of the population residing in cities (4), involving the expansion of infrastructure and shifts in land use patterns and population density. These alterations contribute to increased environmental pollution and decreased availability of natural spaces (4). Climate change results in rising temperatures, changed weather patterns, and extreme weather events (5). These factors are interconnected, and changes in one may trigger or amplify changes in another.

Mental disorders ranked among the three leading causes of health loss globally, consistently contributing to over 14% of age-standardised years lived with disability during the past three decades (6). It has been suggested that adverse macroenvironmental factors contribute to an increased risk of mental health disorders (7–9) and may account for more than 20% of population attributable risk of mental disorders (10,11). While extensive research has explored the influence of the microenvironment on brain and mental health, the significance of the macroenvironment has only recently attracted attention. Mental illness may result from accumulated exposure to single or multiple environmental factors throughout the individual’s life course. In almost all cases, there is a complex interplay between risk and protective factors of micro- and macroenvironment.

In view of these complex dynamics, it is essential to understand how the macroenvironment contributes to the occurrence of mental illness and which are the neurobiological underpinnings of this relationship. In the following sections, we document the association of the macroenvironment with brain structure and function and attempt to connect these findings to potential risks of mental illness. We address the most common macroenvironmental exposures that encompass immediate environmental factors, such as air, noise and light pollution, proximal factors comprising regional socioeconomic characteristics, and distal factors, like urbanisation, natural spaces, and climate. These macroenvironmental exposures are mostly modifiable, presenting opportunities for interventions and strategies to promote the structural and functional integrity of the brain and mitigate the burden of mental illness.

## Search strategy and study selection

We conducted a literature search on the association between modalities of the macroenvironment and magnetic resonance imaging (MRI)-assessed brain structure and function in PubMed from January 1, 2010, to April 19, 2023. We used predefined search terms (**Supplementary Information**), with no restrictions applied except for the filter [Humans]. In short, MeSH (Medical Subject Headings) terms and title/abstract text words related to environmental exposures were employed, including urbanisation, air, noise and light pollution, green space, blue space, regional socioeconomic factors, climate, weather extremes, combined with MRI-detected brain changes in structure and function. The reference lists of relevant systematic reviews identified in our formal search were hand-searched for relevant literature. Furthermore, studies known to the authors were added. Publications on animal models and cell lines were excluded. Studies investigating the association between indoor air pollution and occupational hazards with brain plasticity were further excluded.

## Air pollution

Air pollution arises from natural phenomena, like dust storms or wildfires, and from human activities, such as industrial processes and transportation. It includes solid particles and liquid droplets suspended in the air, referred to as particulate matter (PM), and gases, like ground-level ozone, sulphur dioxide (SO2), nitrogen oxides (NOx: NO + NO2), carbon monoxide (CO), polycyclic aromatic hydrocarbons (PAH) and others (12). Each of these pollutants may have independent and potentially synergistic effects; however, the impact of exposure to a combination of air pollutants on human health is not well understood (13).

Air pollutants enter the body through the respiratory system, initiating a cascade of physiological and biochemical responses affecting different tissues and organs, including the brain (14,15). Pollutants translocate across the blood-brain barrier and can induce systemic inflammation (14), compromising the permeability of blood-brain barrier (16). Air pollution-related neuroinflammation was associated with neurotoxicity, oxidative stress, and impaired control of inflammatory processes (17,18). The developing brain is highly vulnerable to toxicants during two critical developmental periods, the foetal and early life, due to the limited barrier function of the placenta and blood-brain barrier, and potential toxicant transfer during breastfeeding (19).

Prenatal and early childhood exposure to several components of traffic-related air pollution (TRAP), such as PM, PAH, airborne copper and organic carbon, appear to influence brain development in later childhood and adolescence (20–28), including the corpus callosum (21,28), limbic system (21,26), nucleus accumbens (NAc) and cerebellum (21,27) (see **Table 1** for a detailed description). In addition to these structural changes, TRAP exposure is associated with functional connectivity (FC) changes, mainly in frontocortical areas and the default mode network (DMN) (29,30).

**Table 1.** Studies on air pollution and MRI-detected alterations in brain structure and function.

| Exposure period | Exposure duration | Period at MRI assessment | Pollutant/ Method of assessment | Brain alterations | Reference |
| --- | --- | --- | --- | --- | --- |
| Lifetime |  | Childhood to adolescence<br>7 – 18 years | Lived in a highly polluted city vs unpolluted city | Prefrontal WM hypersensitivity<br>↓ WM volumes in temporal and parietal lobe | (227,228)<br>Mexico City and Polotitlán area study<br>n = 30, 73 |
| Childhood<br>9 – 10 years | 1 year | Childhood | PM2.5<br>Spatiotemporal model | ↓ SA in frontal pole (R), cuneus (L)<br>↑ SA in lateral orbitofrontal (R)<br>↓ CT in lateral orbitofrontal (L), superior frontal (L), inferior temporal (R), parahippocampus (R), rostral anterior cingulate (L), caudal anterior cingulate (L), posterior cingulate (L), isthmus (L), insula (R)<br>↑ CT lateral orbitofrontal (R), paracentral (R), middle temporal (L), rostral anterior (R), caudal anterior (R), posterior (R)<br>↓ volumes in accumbens (L), pallidum (R), thalamus (R)<br>↑ volumes in pallidum (L), putamen (L) | (31)<br>ABCD study<br>n = 10,343 |
| Childhood<br>9 – 10 years | 1 year | Childhood | PM2.5<br>Spatiotemporal model | ↑ rN0 (nonlinear) in the cingulum hippocampal portion (L), uncinate fasciculus (L), and fornix (L)<br>↑ rN0 (linear) in the uncinate fasciculus (R), the fornix (R), superior longitudinal fasciculus (L)<br>↓ MD (nonlinear) in the anterior thalamic radiations (L), cingulum hippocampal portion (L), fornix (L),<br>superior longitudinal fasciculus (L), uncinate (L), inferior longitudinal fasciculus (R), and uncinate (R)<br>↓ MD (linear) in the inferior fronto-occipital (L), inferior longitudinal fasciculus (L), cingulum hippocampal portion (R), fornix (R) | (229)<br>ABCD study<br>n = 7,602 |
| Childhood/<br>Preadolescence<br>9 – 13 years | 2 years | Preadolescence/<br>adolescence<br>11 – 15 years | PM2.5<br>Spatiotemporal<br>models | ↑ WM volume in caudate/corpus callosum (L), cingulum (L), inferior fronto-occipital fasciculus, inferior frontal gyrus (R), inferior temporal gyrus (R)<br>↑ GM volume in precentral gyrus (L), cerebellum (L), medial orbitofrontal cortex<br>↓ WM volume in inferior temporal gyrus (L), angular gyrus (L), posterior thalamic radiation (L), middle frontal gyrus (L), hippocampal cingulum (L), postcentral gyrus (R),<br>↓ GM volume in insula (L), cingulate gyrus (R), caudate (R), cerebellum (L), fusiform gyrus, precentral gyrus (R), middle frontal gyrus | (32)<br>San Francisco<br>and San Jose Bay<br>Area study<br>n = 115 |
| Prenatal | Whole pregnancy<br>(PM2.5)<br>48-h during last<br>trimester (PAH) | Childhood/Adolescence<br>6 – 14 years | PM2.5, PAH<br>Spatiotemporal<br>models (PM2.5)<br>Personal air monitors<br>(PAH) | **Exposure to PM2.5**<br>↓ WM surface in lateral pre/procentral gyrus, superior frontal gyrus, middle frontal gyrus (L), middle temporal gyrus (L), inferior parietal lobule (L), anterior cingulate cortex, posterior cingulate cortex (R) with higher exposure to PM2.5<br>↑ WM surface in medial and dorsal pre/procentral gyrus, medial superior frontal gyrus, lateral superior temporal gyrus (R), dorsal superior parietal gyrus with higher exposure to PM2.5 | (26)<br>CCCEH study<br>n = 332 |
|  |  |  |  | ↓ CT in superior parietal gyrus, pre/procentral gyrus with higher exposure to PM2.5<br>↑ CT in superior frontal gyrus, inferior frontal gyrus (L), superior temporal gyrus (L), inferior temporal gyrus, middle temporal gyrus, inferior parietal lobule (L), anterior cingulate cortex, posterior cingulate cortex (R), fusiform and lingual gyrus with higher exposure to PM2.5<br>↑ FA in caudate, lenticular nucleus, insula, brainstem, thalamus, cingulate gyrus, superior corona radiata with higher exposure to PM2.5<br>↑ ADC in inferior fronto-occipital fasciculus, anterior corona radiata, vertical occipital fasciculus with higher exposure to PM2.5<br>**Exposure to PAH**<br>↓ WM surface in inferior temporal gyrus, middle temporal gyrus, inferior parietal lobule (L), anterior cingulate cortex (R), posterior cingulate cortex (L)<br>↑ WM surface in pre/procentral gyrus, superior frontal gyrus, dorsal middle frontal gyrus, ventral fusiform gyrus, ventral lingual gyrus<br>↓ CT in superior frontal gyrus, middle frontal gyrus, inferior frontal gyrus (L), pre/procentral gyrus, superior temporal gyrus (R), middle temporal gyrus (R)<br>↑ CT in middle temporal gyrus (L), anterior cingulate cortex, fusiform and lingual gyrus |  |
|  |  |  |  | ↑ FA in middle orbitofrontal gyrus, cerebellum, hippocampus, globus pallidus, putamen, thalamus, corpus callosum, internal capsule<br>↓ ADC in internal capsule, corpus callosum |  |
| Prenatal | 1<sup>st</sup>, 2<sup>nd</sup>, 3<sup>rd</sup> trimester and whole pregnancy | Childhood/ Preadolescence<br>8 – 12 years | PM2.5<br>LUR models | ↓ volume in total, anterior and body corpus callosum with higher PM2.5 exposure during the 3<sup>rd</sup> trimester. Associations did not survive false discovery rate correction.<br>No association for WM, GM and lateral ventricles | (28)<br>BREATH<br>n = 186 |
| Childhood/ Preadolescence<br>7 – 11 years<br>8 – 12 years | 1 year<br>Two 1-week periods separated by two semesters | Childhood/ Preadolescence | NO2, PAH, BPA, EC, copper<br>Monitors at site | **Exposure to PAH, BPA and NO2**<br>↓ Caudate volume<br>No association for putamen and globus pallidus<br>**Exposure to copper**<br>↑ GM concentration in the caudate nucleus<br>No association for putamen and globus pallidus<br>↑ FA predominantly in caudate nucleus<br>↓ rsFC between the frontal lobe opercula and the caudate nuclei, and vice versa | (29,33,34)<br>BREATH study<br>n ≈ 200 |
| Childhood/ Preadolescence<br>8 – 12 years | 1 year<br>Two 1-week periods separated by two semesters | Childhood/ Preadolescence | Pollution index: weighted average of pooled indoor and outdoor NO2 and EC<br>Monitors at site | ↓ rsFC between regions belonging to the DMN<br>↑ rsFC between the medial frontal cortex and the frontal operculum at the lateral boundary of the DMN | (30)<br>BREATH study<br>n = 263 |
|  |  |  |  | ↓ task fMRI deactivation (rest > task map) in the supplementary motor area and somatosensory cortex |  |
| Prenatal | 1 year | Childhood<br>6 – 10 years | NO<sub>2</sub>, PM<sub>coarse</sub>,<br>PM<sub>2.5</sub>, PM<sub>2.5abs</sub><br>LUR models | ↓ CT in praecuneus (R), pars opercularis (R), pars orbitalis (R), rostral middle frontal (R), superior frontal (R), cuneus (L) with higher exposure to PM<sub>2.5</sub><br>↓ CT in lateral orbitofrontal (R) with higher exposure to PM<sub>2.5coarse</sub><br>↓ CT in fusiform (L) with higher exposure to PM<sub>2.5abs</sub> | (21)<br>Generation R<br>study<br>n = 783 |
| Prenatal | 1 year | Preadolescence<br>9 – 12 years | NO<sub>x</sub>, NO<sub>2</sub>, PM<sub>10</sub>,<br>PM<sub>coarse</sub>, PM<sub>2.5</sub>,<br>PM<sub>2.5abs</sub>, PAH, OC,<br>copper, iron, silicon,<br>zinc, OP<br>LUR models | ↑ volumes of putamen and pallidum with higher exposure to PM<sub>coarse</sub><br>↑ volume of cerebellum with higher exposure to PM<sub>10</sub>, PM<sub>coarse</sub>, PM<sub>2.5</sub>, PM<sub>2.5ab</sub><br>↓ volume of hippocampus with higher exposure to PAH, copper<br>↓ volume of amygdala with higher exposure to OC, silicon<br>↓ volume of corpus callosum with higher exposure to OP<br>↓ CT in postcentral gyrus with higher exposure to OC | (21)<br>Generation R<br>study<br>n = 3,133 |
|  |  |  |  | ↓ CT in rostral middle frontal gyrus with higher exposure to copper and PM2.5abs<br>↓ FA in forceps minor, corticospinal tract, superior longitudinal fasciculus (R) with higher exposure to PM2.5<br>↑ MD in cingulum bundle, forceps minor, superior longitudinal fasciculus (L), inferior longitudinal fasciculus (L) with higher exposure to silicon<br>↑ rsFC between brain regions of the same brain hemisphere, predominantly in the auditory association, dorsolateral prefrontal, somatosensory and motor, anterior cingulate and medial prefrontal, dorsal stream visual, and insular and frontal opercular cortices with higher exposure to NO2 |  |
| Childhood<br>6 – 10 years | 1 year | Preadolescence<br>9 – 12 years | NOx, NO2, PM10, PMcoarse, PM2.5, PM2.5abs, PAH, OC, copper, iron, silicon, zinc, OP<br>LUR models | ↓ volume of hippocampus with higher exposure to PMcoarse, OP<br>↑ volume of nucleus accumbens with higher exposure to zinc<br>↓ volume of corpus callosum with higher exposure to OC<br>↓ CT in lingual gyrus with higher exposure to copper and OP<br>↑ SA in precentral gyrus with higher exposure to zinc and OP | (21)<br>Generation R<br>study<br>n ≈ 3,000 |
|  |  |  |  | ↑ SA in pericalcarine cortex and precuneus with higher exposure to zinc<br>↓ SA in pars triangularis with higher exposure to PMcoarse<br>↓ FA in corticospinal tract (L), uncinated fasciculus, superior longitudinal fasciculus (R), inferior longitudinal fasciculus (R) with higher exposure to NOx<br>↑ MD in cingulum bundle (L) with higher exposure to OP<br>↑ MD in cingulum bundle, forceps minor, superior longitudinal fasciculus, inferior longitudinal fasciculus, uncinated fasciculus with higher exposure to zinc |  |
| Prenatal and childhood | Whole pregnancy and childhood at periods:<br>0 – 2 years<br>2 – 5 years<br>5 – 9 years | Preadolescence<br>9 – 12 years | NOx, NO2, PM10, PMcoarse, PM2.5, PM2.5abs<br>LUR models | **Exposure to NOx from 0 – 2 and 2 – 5 years**<br>↑ rsFC in areas in auditory association, premotor, orbital and polar frontal, inferior parietal, and posterior cingulate cortices, in the ventral diencephalon, and in the MT+ complex and neighbouring visual areas<br>**Exposure to NO2 during pregnancy and from 0 – 2years**<br>↑ rsFC in areas in in auditory association, ventral diencephalon, and insular and frontal cortices opercular, somatosensory and motor and early auditory, dorsal stream visual and superior parietal<br>**Exposure to PMcoarse from 2 – 5 and 5 – 9 years** | (21)<br>Generation R study<br>n = 2,197 |
|  |  |  |  | ↑ rsFC in areas in anterior cingulate and medial prefrontal cortices and in the MT+ complex and neighbouring visual areas<br>**Exposure to PM2.5abs from 0 – 2 and 2 – 5 years**<br>↑ rsFC in areas in insular and frontal opercular, auditory association, lateral temporal, somatosensory and motor, anterior cingulate and medial prefrontal, and posterior cingulate cortices, and in the MT+ complex and neighboring visual areas |  |
| Prenatal and childhood | Whole pregnancy and childhood at periods<br>0 – 3 years<br>3 – 6 years<br>6 – age of MRI | Preadolescence<br>9 – 12 years | NOx, NO2, PM10, PM2.5, PM2.5abs<br>LUR models | **Exposure to NOx from 3 – 6 years**<br>↑ rsFC in regions of the visual and task positive networks: MT+ complex and neighbouring visual areas – inferior frontal cortex and MT+ complex and neighbouring visual areas – insular and frontal opercular cortex<br>**Exposure to NO2 from 0 – 3 years**<br>↑ rsFC in regions of the visual, auditory and task positive networks: dorsal stream visual cortex – superior parietal cortex and auditory association cortex – insular and frontal opercular cortex<br>**Exposure to PM2.5abs from 0 – 3 years**<br>↑ rsFC between brain regions of several networks (19 of 22): visual - visual, visual – auditory, visual – task positive, visual – task negative, auditory – task positive, auditory – task negative, and task negative – task negative<br>↓ rsFC between brain regions of visual – task positive networks and task positive – task negative networks (3 of 22): MT + complex and neighbouring visual areas – superior parietal cortex, posterior<br>cingulate cortex – superior parietal cortex, and frontal opercular cortex – lateral temporal cortex | (75)<br>Generation R study<br>n = 2,197 |
| Prenatal and childhood | 48-h during last trimester<br>5 years of age | Childhood<br>Mean age ±SD:<br>8.0 ±1.3 years | PAH<br>Personal air monitors during last trimester<br>Urine samples in childhood | **Prenatal exposure to PAH**<br>↓ local volume in the middle frontal gyrus, medial orbitofrontal gyrus, inferior frontal gyrus, superior frontal gyrus, pre-central gyrus, post-central gyrus, supramarginal gyrus, middle temporal gyrus, superior temporal gyrus, mesial superior parietal gyrus, praecuneus, cuneus, cingulate gyrus, gyrus rectus in the left hemisphere.<br>No association with cortical thickness<br>**Postnatal exposure to PAH**<br>↓ WM surface in dorsolateral prefrontal regions, especially over the superior frontal gyri | (25)<br>CCCEH study<br>n = 40 |
| Lifetime | From birth to 12 years of age | Childhood<br>12 years | EC (high vs low exposure group)<br>LUR models | ↓ CT in the medial frontal gyrus , ventromedial prefrontal cortex (R), paracentral lobule, postcentral gyrus, superior frontal gyrus, precentral gyrus (L), inferior parietal lobule (L), superior parietal lobule (L), anterior cingulate (L), cingulate (R), precuneus (L), fusiform gyrus (L)<br>↓ GM volume in the cerebellum, precentral gyrus, inferior parietal lobule | (27)<br>CCAAPS study<br>n = 135 |
| Adulthood | 1 year | Adulthood<br>≥ 60 years<br>Median age [IQR]:<br>68.0 [9.0] years | PM2.5<br>Spatiotemporal model | ↓ total cerebral brain volume<br>No association with hippocampal volume | (38)<br>Framingham Offspring Study<br>n = 943 |
| Adulthood | 1 year | Adulthood<br>44 – 80 years | NOx, NO2, PM10, PMcoarse, PM2.5<br>LUR models | ↓ total GM volume with higher exposure to any of the investigated pollutants<br>↓ GM volume in the frontal pole and operculum cortex (L) with higher exposure to PM10 | (39,47,49)<br>UK Biobank<br>n ≈ 18,290 |
|  |  |  |  | ↓ GM volume in the frontal pole, operculum cortex (L), and orbital cortex (R) with higher exposure to NO<sub>x</sub><br>↓ GM volume in the frontal pole (R) and operculum cortex (L) with higher exposure to NO<sub>2</sub><br>**Exposure to PM<sub>2.5</sub>**<br>↓ total WM volume<br>↓ GM volume in the frontal pole, orbital cortex (R), operculum cortex (L)<br>**Exposure to PM<sub>coarse</sub>**<br>↓ GM volume in the frontal pole (R), superior gyrus (L), operculum cortex (L)<br>↓ volume in the thalamus (L) |  |
| Adulthood | 3, 8, and 10 years | Adulthood<br>71 – 89 years | NO<sub>2</sub>, PM<sub>2.5</sub>, diesel<br>PM<br>Spatiotemporal model | **3-year cumulative exposure to NO<sub>2</sub>**<br>↓ GM volumes in the prefrontal cortex<br>↓ volumes in the anterior cingulate gyrus, insula, amygdala, limbic medial temporal lobe, basal ganglia<br>**3-year cumulative exposure to PM<sub>2.5</sub>**<br>↓ WM volumes in the anterior and posterior extreme/ external capsule, calcarine gyri<br>↓ GM volumes in the superior, middle, medial frontal gyri, inferior frontal gyrus (L), superior parietal lobule, occipital poles | (40–42,48,50)<br>WHIMS<br>n = 764, 1,403, 1,365<br>n = 1,403 (8- and 10-year assessment) |
|  |  |  |  | ↓ volume in the anterior cingulate gyrus<br>↑ volumes in the thalamus, putamen, globus pallidus, posterior insula<br>No association with volumes of corpus callosum, hippocampus, temporal lobe<br>**8-year cumulative exposure to PM2.5**<br>↓ total WM volume and in the frontal, parietal, temporal lobes, corpus callosum<br>No association with hippocampal, basal ganglia volumes and GM volumes across the cerebral cortex<br>**10-year cumulative exposure to diesel PM**<br>↑ ventricular volume<br>U-shaped associations were observed for total WM volumes and in frontal, parietal and temporal lobes<br>↓ total GM volumes and in frontal, parietal and temporal lobes |  |
| Adulthood | 5 – 20 years divided in 8-year periods | Adulthood<br>Mean age ±SD:<br>76.0 ±5.0 years | PM10, PM2.5<br>Spatiotemporal model | ↓ deep-GM volumes<br>↓ volumes in total brain, frontal and parietal lobe in one of the study centres with higher exposure to PM2.5<br>No association with hippocampal volume | (43)<br>ARIC study<br>n = 1,753 |
| Adulthood<br>50 – 80 years | 1 year | Adulthood<br>55 – 85 years | NOx, NO2, PM10,<br>PM2.5, PM2.5abs<br>LUR models | Local atrophy in inferior parietal lobule (R) with higher exposure to NOx, PM10, and PM2.5 | (44,52)<br>1000BRAINS<br>n ≈ 600 |
|  |  |  |  | Local atrophy in posterior cingulate cortex and praecuneus (R) with higher exposure to NOx, NO2, and PM10<br>No association with local atrophy in the dorsolateral prefrontal cortex<br>↓ Intra-network rsFC and segregation index in the dorsal attention network with higher exposure to NO2<br>↓ Intra-network rsFC in the ventral attention network with higher exposure to PM10 and PM2.5<br>↑ Inter-network rsFC in the visual network with higher exposure to PMabs<br>↓ segregation index in the ventral attention network with higher exposure to PMabs |  |
| Adulthood | 1 year | Adulthood<br>Mean age ±SD:<br>49.5 ± 13.3 years | NO2, PM2.5, ozone<br>LUR models | ↑ volume in rostral middle frontal (L), supramarginal (L), transverse temporal (L), pars opercularis (R) with higher exposure to NO2<br>↑ volume in pars triangularis (L) and CT in fusiform (R) with higher exposure to PM2.5<br>↑ pars orbitalis volume (L) with higher exposure to ozone<br>No association with WM and GM volumes | (45)<br>Taiwanese sleep study<br>n = 4,866 |
| Adulthood | 5 years (NO2, PM10)<br>1 year (PM2.5) | Adulthood<br>Mean age ±SD:<br>67.3 ± 6.4 years | NO2, PM10, PM2.5, PAH metabolites<br>LUR models | **Exposure to NO2**<br>↓ volume in caudate, pallidum, amygdala, nucleus accumbens | (46,230)<br>EPINEF study<br>n = 957<br>n = 528 (PAH) |
|  |  |  | Urine samples (PAH) | ↓ CT in the frontal cortex, lateral temporal cortex, inferior parietal cortex, posterior cingulate, insula, parahippocampal gyri, fusiform gyri<br>↑ CT occipital cortex, postcentral gyri (L)<br>**Exposure to PM10**<br>↓ volume in pallidum, putamen, amygdala, nucleus accumbens<br>↓ CT in the lateral temporal cortex, inferior parietal cortex, prefrontal cortex, posterior cingulate, insula, parahippocampal gyri, fusiform gyri<br>↑ CT occipital cortex, postcentral gyri<br>**Exposure to PM2.5**<br>↓ volume in nucleus accumbens<br>↓ CT in the lateral temporal cortex, inferior parietal cortex, prefrontal cortex, insula, parahippocampal gyri, fusiform gyri<br>↑ CT occipital cortex, postcentral gyri<br>**Exposure to PAH (highest vs lowest quartile)**<br>↓ CT in parietal, temporal and insular lobes in men<br>↓ CT in frontal and parietal lobes in women<br>↓ volumes in the caudate in men, and pallidum in women |  |
| Adulthood | 120 min | Adulthood<br>Mean age ±SD:<br>27.4 ±5.5 years | Diesel exhaust<br>(intervention arm) | No differences in the default mode network rsFC for post- compared to pre-diesel exhaust exposure | Randomised cross-over study (231) |
|  |  |  | Filtered air (control arm) | ↑ rsFC in the middle temporal gyrus (R), occipital fusiform gyrus (R) for post-filtered air compared to pre-filtered air<br>↑ rsFC in the angular gyrus (R), frontal pole, middle frontal gyrus, middle temporal gyrus, praecuneus cortex, temporal pole (L) for post-filtered air compared to post-diesel; exhaust | n = 25 |

Abbreviations: ABCD, Adolescent Brain Cognitive Development; ADC, average diffusivity coefficient; ARIC, Atherosclerosis Risk in Communities; BPA, benzo[a]pyrene; CCAAPS, Cincinnati Childhood Allergy and Air Pollution Study; CCCEH, Centre for Climate Change and Environmental Health; CT, cortical thickness; EC, elemental carbon; EPINEF, Environmental Pollution-Induced Neurological Effects; FA, fractional anisotropy; FC, functional connectivity; GM, gray matter; L, left; LUR, land-use regression; MD, mean diffusivity; MRI, magnetic resonance imaging; NO<sub>x</sub>, nitrogen oxides; NO<sub>2</sub>, nitrogen dioxide; OC, organic carbon; OP, oxidative potential of PM<sub>2.5</sub>; PAH, polycyclic aromatic hydrocarbons; PM, particulate matter; PM<sub>2.5</sub>, particulate matter with aerodynamic diameter ≤2.5 μm; PM<sub>2.5</sub>abs, absorbance of the PM<sub>2.5</sub> fraction; PM<sub>10</sub>, particulate matter with aerodynamic diameter ≤10 μm; PMcoarse, particulate matter with aerodynamic diameter between 10 μm to 2.5 μm; R, right; rNO, restricted isotropic intracellular diffusion; rs, resting state; SA, surface area; WHIMS-MRI, Women's Health Initiative Memory Study-MRI; WM, white matter;

Prenatal exposure to fine PM (PM2.5) and PAH is associated with smaller white matter (WM) volume in parietal lobes (26), and WM surface reductions in the left hemisphere, mediating the association between air pollutants and conduct disorder problems (25). Furthermore, early life exposure to TRAP is associated with increased frontotemporal cortical thickness in children and adolescents (21,26,27). Alterations in global WM microstructure, including increase fractional anisotropy, and in several WM microstructure tracts were documented (21). Hemispheric asymmetry in WM and gray matter (GM) volumes across all cortical regions and several subcortical regions has been observed (29,31–34). While brain asymmetry is a typical trait in humans, it can be altered and has been linked to psychiatric disorders (35–37).

The vulnerability of the brain to air pollution extends beyond early brain development and includes later stages of life. Studies among adults exposed to different components of air pollution, reported volume reductions in total cerebral brain (38), total WM and GM (39–42), deep-GM (43) and local atrophy mainly in frontocortical areas, insula and subcortical regions (44–51), which partially mediated the association between PM2.5 and NO2 with depressive symptoms (50).

Functional neuroimaging studies reported a reduced stress-related activation in connectivity networks associated with acute stress, such as the salience, DMN, and central executive networks, in adults with higher exposure to air pollution (52), and augmented stress-related information transfer across cortical and subcortical brain networks among participants with a higher polygenic risk score for depression (53); suggesting that air pollution may increase vulnerability to mood dysfunction and potentially inhibit an appropriate stress response.

Taken together, it becomes apparent that exposure to air pollution has diverse and hemisphere-specific implications on brain morphology and function in children and adults (**Table 1**). Air pollution effects on brain regions appear to vary depending on the specific pollutant and period of assessment during the lifespan. Although concrete conclusions cannot be made, disruptions were observed in regions such as prefrontal cortex (PFC), ACC, hippocampus, amygdala, insula, NAc, corpus callosum and striatum, all of which have been implicated in the risk for major psychiatric disorders (54,55), like depression, anxiety (56–59), substance use disorders (60,61) and schizophrenia (54,62).

Epidemiological studies have provided evidence linking air pollution to mental health disorders in exposed youth and adults (63). Recent meta-analysis highlighted a positive association between PM2.5, PM10 and NO2 exposure with risk for depression (72) and suicide (64). Furthermore, evidence supports that short- and long-term exposure to PM2.5 is linked to an increased risk for anxiety, while exposure to PM10, NO2, and NOx might increase the risk for schizophrenia or hospitalisation for schizophrenia (65,66). By linking epidemiological approaches on air pollution with neuroimaging data, future studies can help elucidate mechanisms by which air pollution-induced neuroinflammation and other potential biological pathways, such as stress response (17) may affect brain, behaviour and psychopathology.

## Noise pollution

Noise pollution originates from urban traffic, airports, industries, and construction sites and can evoke negative emotions and annoyance. Prolonged exposure to disruptive noise is thought to induce brain alterations through mechanisms such as sleep disturbances, which prompt a pro-oxidative environment, predisposing to neuroinflammation, and heightened hypothalamic-pituitary-adrenal (HPA)-axis reactivity (67,68), that might contribute to mental illness (69,70). Residents in a community impacted by changed flight patterns compared to a demographically similar non-impacted community, showed a higher risk for substance use and mental health-related emergency visits among individuals living in noise-affected communities, particularly in younger age groups (71). Meta-analyses have reported increased odds for depression and anxiety with higher 24-h noise level (72). Still, the association between noise and mental health is limited due to high risk-of-bias studies and inconsistent findings across studies included in the different systematic reviews (72–74).

The relation between noise pollution and brain structure and function remains understudied and is also afflicted with inconsistent findings (75,76). A study on 8–12 year-olds exposed to school road-traffic noise over one year reported enhanced connectivity in the subcortical auditory pathway (77), indicating possible enhancement on auditory processing abilities but also increased sound sensitivity and sensory overload. Whether these results, along with potential noise-induced chronic stress and sleep disturbance contribute to anxiety and behavioural problems in children requires further investigations. Among older adults participating in a 5-year study, higher noise pollution was associated with cognitive decline and alterations in brain network organisation were found (44,52). Further studies on the behavioural and cognitive consequences of noise pollution across the lifespan are required to provide robust evidence and establish explicit mediating brain structures and functions.

## Light Pollution

Light pollution, a consequence of human activities, including outdoor lighting, commercial signage, and illuminated buildings, produces excessive or misdirected artificial light and disrupts the natural darkness of the night sky. Exposure to artificial light at night (ALAN) has become increasingly prevalent, especially in urban areas. Light is detected by the retina and transmitted through the intrinsically photosensitive retinal ganglion cells (ipRGCs) to the suprachiasmatic nucleus in the hypothalamus and other brain regions involved in regulating circadian rhythms and sleep-wake cycles (78). Circadian rhythm disruptions have been linked to an elevated risk of major depressive disorders, bipolar disorders, and heightened mood instability (79), potentially mediated by oscillations in clock genes expression responsive to light-dark transitions (80). Light is also projected (via the ipRGCs and the suprachiasmatic nucleus) to regions involved in mood regulation, such as the PFC, hippocampus, and amygdala (81,82), directly influencing emotional processing and mood functions (83,84). Hence, prolonged and ill-timed ALAN exposures may precipitate or worsen symptoms of mood disorders.

Cross-sectional analyses reported an increased prevalence of mood and anxiety disorders in adults and adolescents with higher exposure to outdoor ALAN (85–87). However, residual confounding due to air pollution has likely influenced the results (87). We found no studies examining the relationship between ALAN and brain structure and function. Participants exposed to dim ALAN during one-night sleep in a polysomnography laboratory exhibited decreased brain activity in the inferior frontal gyrus (IFG) compared to a night without any light exposure (88). Decreased activation in the IFG has been associated with impairments in executive functions and reported in clinical populations afflicted with bipolar disorder, depression, and schizophrenia (89–92). Still, further research is needed to elucidate the effects of light pollution on brain changes.

## Urbanisation

Urbanization is a shared element in global migration patterns over the past half-century, involving the transition from rural to urban settlements (4). Historically, this transition has been linked to economic growth. Urban dwellers are more likely to benefit from sustainable infrastructure, essential education, healthcare services, and more work opportunities than rural residents. Despite these advantages, urban environment is inhomogeneous, depicted by economic, social and environmental inequalities (4,93). Rapid and unplanned urbanisation increases income inequalities, linked to disparities in health and education, marginalisation, social isolation and threat, and environmental pollutants (93–95). The urban environment is associated with mental disorders, such as depression, anxiety and schizophrenia (95–100), with urban upbringing identified as the most prominent risk factor for schizophrenia (9,98).

A common underlying mechanism linking urban living stressors to vulnerability to mental illness has been suggested to be the dysregulation of the HPA-axis (17,93,101,102), potentially resulting in cerebral functional and structural changes (103). Moreover, urban environments may interact with genetic variations in genes related to stress response and brain structure, such as neurodegeneration, neural differentiation, and axon growth (104). Various neuroimaging studies reported the association between urban environment and functional changes in stress-related brain regions (105,106). Current city living and urban upbringing associated with increased activity in the amygdala-hippocampus complex and subgenual ACC during a stress task in healthy adults (107,108). The ACC is a key region for regulating amygdala function, negative emotions, and stress and has been proposed to mediate the relationship between medial PFC (mPFC) activity and affective symptoms (109). Among individuals with an urban upbringing, activity alterations in those brain regions were modulated by genes related to dopamine, anxiety, and stress phenotypes, whereas such effects were not evident in individuals with a rural or small-town childhood (110,111).

Urban upbringing is associated with reduced hippocampal and amygdala volumes among adolescents (112) and dorsolateral PFC (dlPFC) and mPFC in adults (113,114). Healthy adults with higher urban upbringing scores were observed to have cortical thinning in the dorsolateral, mPFC, and parahippocampal cortex (115), although findings are not consistent (116). Stress-induced volume reductions in the observed regions during childhood are associated with depression, psychosis, and post-traumatic stress disorders in later life (117–119), while the identified cortical thinning aligns with regions implicated in psychiatric conditions, including schizophrenia and bipolar disorders (120). To what extent brain changes in these disorders are driven by urbanicity remains to be determined.

The urban environment encompasses various economic, social, ambient, and infrastructural characteristics. Current literature assessed urban living based on a measure of population density and duration of residency (literature is described in this section) or by using isolated factors, such as pollution, urban green spaces, and socioeconomic deprivation, which often co-occur and interact within individuals’ living environment. A study using a composite measure of urban living, including night-time lights, green space, build-up space, water bodies and land use, reported an association with reduced mPFC volume, increased cerebellum volume, and decreased functional network connectivity within the mPFC of the anterior DMN that was observable in two cohorts of young adults residing in Europe and China. The observed neural correlates mediated the association between urban living and depressive symptoms (121). In addition, analyses on a comprehensive set of factors related to urban living identified environmental profiles relevant to psychiatric symptoms among adults. In particular, an environmental profile predominantly characterised by regional deprivation, pollution and density of urban infrastructure was positively associated with affective symptoms and mediated by smaller striatum volumes, while an environment characterised by dense build-up space and mixed land use was associated with anxiety symptoms and was mediated by reduced volumes of IFG, amygdala and cerebellar regions. The associations were moderated by genes related to stress response regulation, anxiety and phobia, suggesting that genetic variations may explain individual differences in response to environmental adversity (104).

Further research is warranted that accounts for the inherent complexity of the living environment to disentangle the distinct and interconnected attributes of urban environments that contribute to brain function and dysfunction.

## Natural space

Two prominent frameworks have been suggested to explain the effects of natural environments, such as surrounding green spaces, forests, or water bodies on mental well-being. The attention restoration theory posits that nature facilitates the restoration of attentional capacity, reduces mental fatigue, and enhances cognitive functioning (106,122–124). Simultaneously, the stress reduction theory proposes that nature lowers stress levels and enhances positive feelings (125,126). These effects occur via mechanisms involving the autonomic nervous system, reflected by lower blood pressure and improved heart rate, as well as the modulation of the endocrine system, including reductions in stress hormones secretion (127,128). Nature-induced benefits on the central nervous system have also been observed in experimental, intervention and observational studies, corroborating the notion that contact with nature promotes mental health. Compared to urban scenes, viewing natural landscapes in a laboratory setting was linked with cognitive restoration, reduced visual attention focus (129), and activation of brain areas associated with positive emotional responses, rewarding experience, and recollection of positive memories (105,130–132). Additionally, nature images evoked enhanced FC between the DMN, dorsal attention network, ventral attention network, and the somatomotor network, potentially promoting cognitive coherence and effortless attentional engagement (133).

Walking in nature showed positive effects on brain and mental health by decreasing PFC activation, which is associated with sadness and behavioural withdrawal, and reducing rumination – a pattern linked to depression (134), possibly via restorative benefits of nature. Additionally, after a nature walk, there was decreased amygdala activation during a social stress-inducing task, a region responding to fear and stress (135). Such benefits were not observed after urban walks (134,135). Surrounding urban green space also seemed to have supportive effects on coping with stress (aligning with the stress reduction theory), as indicated by activation patterns in emotion-regulatory brain areas, like the dlPFC, mPFC, insula, ACC, posterior cingulate, and ventral striatum (136,137). Nevertheless, opposite activation directions than expected were observed in the amygdala that could not be explained with certainty (137).

Higher residential greenness was further associated with morphological brain changes. The findings encompassed higher GM and WM volumes in PFC and cerebellum (39), and lower global atrophy and thicker PFC, insula and praecuneus in adults (138–140) – structures linked to cognitive process and psychiatric disorders when reduced (141–144). Indeed, reduced volumes in frontolimbic and cerebellar regions were observed in environments characterised by reduced access to natural spaces that mediated the association between urban living and affective and anxiety symptoms (104). Further research is needed regarding the different typologies of natural spaces and vegetation, which is currently lacking. For example, among older adults, only neighbourhood forest exposure seemed to positively affect amygdala integrity, but not urban green or blue spaces (145). The presence of green space in the living environment was associated with reduced risk of depression and anxiety in cross-sectional studies (146), however, non-consistently, and similar associations were not supported by longitudinal studies (97,147). These discrepancies possibly arose due to methodological shortcomings, such as an inability to assess whether participants spent time in those environments, and the mediating effects of air and noise pollution or exercise uptake. Different buffer areas around the home location were used in the literature. Yet, it remains unclear which catchment scale is the most relevant for mental wellbeing. Furthermore, distance to green areas was typically calculated with Euclidean distance rather than network or road connections. This approach may not accurately reflect the experiences of the local urban population.

## Regional socioeconomic status

Regional socioeconomic status can significantly influence the cognitive, emotional, and behavioural development of children and adolescents, and these effects may persist in adulthood (148–151). Youth growing up in disadvantaged neighbourhoods, marked by poverty, violence, poor housing conditions, or limited access to educational and healthcare resources (152), are often exposed to higher levels of chronic stress and unpredictability (153), and may have difficulties building supportive social networks (154). Consequently, they face a higher risk of childhood mental disorders (155,156). Neighbourhood disadvantage has been linked to HPA-axis dysregulation and reactivity (157,158), and alterations in neural development and functioning related to cognitive processes, rewards, and social threats in youth. For instance, lower neighbourhood socioeconomic status associated with decreased activation in regions of the executive system, including the dlPFC, posterior parietal cortex, praecuneus and cerebellum, during a working memory task (159). Neighbourhood poverty may also disrupt self-control development, measured with inhibition performance task, via its effect on IFG activation (160). Furthermore, youth living in more deprived areas recorded lower activation in caudate, putamen, accumbens area and pallidum during reward anticipation (161), and higher amygdala reactivity to threat-related stimuli, particularly in neighbourhoods where safety and management norms were more permissive (162). Altogether, these responses have been implicated in internalising and externalising symptoms and psychopathology (163–165).

Changes in connectome in youth residing in socioeconomically disadvantaged areas suggest that neighbourhood deprivation impede the developmental progression of brain function in children and young adults (172), involving reduced fronto-amygdala and fronto-striatal resting state FC (173–175), and changes in FC between DMN and dorsal attention network and sensorimotor systems (166). The observed connectome alterations were coupled with internalising symptoms and worse cognition. Furthermore, increased fronto-striatal FC in newborns living in deprived neighbourhoods mediated the relationship between disadvantage and externalising symptoms at age 2 years (167).

Compared to the above findings, different patterns in FC were observed when community violence and crime were assessed. Such experiences associated with FC changes in youth between regions of the limbic system, mainly encompassing the hippocampus (178,179). Furthermore, youth exposed to community violence demonstrated FC changes between the hippocampus and insula, with opposing directions observed across studies (168,169). These discrepancies may be influenced by various factors, including the specific timing of exposure to community violence, developmental changes, individual characteristics, or other contextual factors, such as positive parenting and school environment (166,170,171). Differential social experiences, such as discrimination, within similar environments may exert distinct neural influences on minoritized and discriminated individuals, including various racial and gender identities, particularly in the domains of threat, reward and emotional processes (153,172–174).

Neighbourhood adversity in adolescence may shape neural responses to social situations, threats, and rewards in adulthood. Individuals with a disadvantaged upbringing displayed increased sensitivity in reward-related brain regions like the striatum, NAc, and ventrolateral PFC. Notably, current income did not mediate the observed associations, suggesting a potential link between early experiences and reward anticipation and pursuit in later life (175). Furthermore, exposure to neighbourhood disadvantage during adolescence might influence reward-related processes in adulthood, via decreased activation in brain regions associated with cognitive and affective processes, such as amygdala, hippocampal and dlPFC (176). Lastly, neighbourhood quality might influence neural responses to social stimuli, as observed by increased activity in the dorsal ACC and prefrontal regions among individuals with disadvantaged upbringing (177).

Several studies have demonstrated the effect of neighbourhood disadvantage on brain structure in youth and adults, such as widespread lower volume of WM and GM (178,179), including hippocampus (168,179–181), amygdala (168), dlPFC and dorsomedial PFC, superior frontal gyrus (181), IFG and ACC (182). In addition, smaller surface area and cortical thinning was observed in the frontal, parietal, and temporal lobes, cingulate and insula (183–187). Finally, neighbourhood disadvantage was linked to atypical neurodevelopmental trajectories during adolescence, indicating delayed brain development (188,189). It is currently unknown whether the deviations in brain trajectories due to adversity, converge later in development or if they reflect atypical developmental patterns.

Altogether, neighbourhood disadvantage was associated with alterations in brain regions involved in emotional processes, including the amygdala, hippocampus, and dlPFC, and reward-related regions such as the striatum and NAc. Several studies accounted for individual or family socioeconomic status as a confounding variable, demonstrating that regional socioeconomic status may exert distinct effects on brain and behaviour. Most studies evaluated neighbourhood disadvantage as a single measure of neighbourhood violence or poverty, or used a composite score structured from several measures, e.g., poverty, unemployment rate, education levels. However, assessing different attributes of regional challenges might elucidate distinct neural correlates to different adversity typologies (190).

## Weather patterns and climate change

Weather patterns encompass various meteorological factors, including temperature, precipitation, humidity and sunlight duration. <u>Mounting evidence suggests that weather patterns may influence mood, behaviour, and overall mental well-being</u>. Higher ambient temperatures have been associated with an increased suicide or self-harm burden (64,191,192), mental health-related mortality, and morbidity of schizophrenia, mood disorders, and anxiety disorders (193,194). Likewise, higher humidity has been linked with a greater burden of concurrent depression and anxiety, increased mental health-related emergency visits (194,195), and aggravation of the adverse effects of high temperatures (196). Regarding precipitation patterns, limited evidence suggests a possible positive link with mental illness (197–199). Studies have reported a negative association between sunlight exposure and risk of depression and anxiety (146), while cloudiness and decreased sunshine duration were linked to increased suicide rates (200). Furthermore, seasonal changes directly affect the duration of daylight. Seasonal daylength fluctuations appear to affect mood and behaviour negatively and were associated with a higher prevalence of seasonal affective disorder and earlier onset of bipolar disorder (201). Here it is important to acknowledge that many of these findings are susceptible to bias due to inadequate control of confounders and the risk for an ecological fallacy – the incorrect inference about individuals based on aggregated-level data associations (202).

The changes in weather patterns associated with climate change introduce new challenges that further complicate mental health outcomes via direct effects of stress and trauma and indirect mediating factors, including food insecurity, poverty, climate change-induced violence and forced migration (100). Extreme weather events include heatwaves, flooding, and drought. Systematic investigations demonstrated positive associations between heatwaves and mental health-related morbidity (193,203), where greater frequency, duration, and intensity of heatwave conditions magnified the observed effects (193,204). Direct exposure to floods was associated with depression, anxiety, post-traumatic stress disorder, suicidal ideation, and psychological distress (194,205,206). Similarly, droughts were associated with increased psychological distress, especially among rural inhabitants and vulnerable populations (207,208). The neural circuits linking weather and psychiatric risk are unclear, as studies investigating the weather and climate change effects on MRI-detected brain activity are lacking. During simulated hyperthermia conditions (50°C, >40min), there was heightened activation in the dlPFC and the right intra-parietal sulcus (209). Additionally, impairments in the FC of the DMN were observed (210), coupled with prolonged reaction time in cognitive tasks compared with the control group (209,210). A few cross-sectional studies reported positive associations between daylength and volumes of the hippocampus, amygdala, and brain-stem – regions that exhibited seasonal variations in serotonin signalling (201), suggesting that changes in volumes of subcortical regions and neurotransmitter signalling involved in emotional regulation may be involved in the seasonality of mental disorders.

## Perspective

The existing literature supports the notion that macroenvironment can influence the physiological development and ageing of the brain. However, reaching definitive conclusions is challenging. Current findings are either contradicting or lack specificity, as multiple regions show an association with macroenvironmental adversity, particularly in relation to air pollution. These observations may result from the diverse selection of regions of interest, the timing and severity of exposure. The influences of macroenvironmental adversity on the brain may be more immediate or manifest over time depending on the specific exposure and brain region (20,121), while the reversibility of unfavourable changes in structure and function following exposure to factors that contribute to resilience is unclear (211).

Research investigating the associations of light and noise pollution, weather patterns and extremes on the brain is notably limited. Certain brain regions have been consistently reported to show changes in response to the other macroenvironmental factors. The common brain areas include regions involved in emotional regulation, such as PFC, amygdala, hippocampus, and ACC, similar to the effects observed in microenvironmental adversity (Vaidya et al., 2023), as well as regions related to reward processing, such as striatum and NAc. More specifically, urbanicity, air pollution, and regional deprivation demonstrated unfavourable effects on these brain regions, while natural spaces were associated with beneficial effects. Furthermore, distinct neural regions have also been associated with different types of environmental adversity. For example, current city living was associated with amygdala activity, while urban upbringing affected ACC (108). Similarly, neighbourhood poverty appeared to impact FC between PFC, amygdala, and striatum, while changes in FC mainly involving the hippocampus were observed with exposure to neighbourhood violence. The underlying pathways for the differential links of various macroenvironmental factors with specific brain regions, despite eliciting common effects, such as HPA-axis activation and neuroinflammation, need to be investigated.

## Future research directions

### Overcoming the complexity of high dimensional data

The associations between the macroenvironment, brain outcomes and mental health involve complex interactions between multiple environmental exposures, individual susceptibility and social factors. As individuals are typically exposed to multiple stressors simultaneously, it becomes challenging to quantify the impact of a specific environmental factor. Furthermore, high correlations are usually present among the different environmental factors adding complexity in determining their independent effects. High collinearity might lead to unstable or imprecise coefficient estimates (212). Indeed, most studies have primarily focused on exploring the relationships between a singular exposure and brain outcomes or mental health, while investigations incorporating analyses of multiple exposures have shown that associations observed with single exposures tend to be less pronounced (21). A further constraint in the existing literature, which impedes the understanding of precise mechanisms, is the insufficient investigation into the mediating role of brain structure and function in the association between the environment and mental health.

To address these challenges, statistical models are needed that enable simultaneous modelling of high-dimensional data, aiming to reduce the complexity and understand underlying patterns by grouping them based on their shared characteristics and distinctions. These methods include independent component analysis, canonical correlation analysis, hierarchical clustering, latent class analyses, and normative modelling (213). An example of such analyses has been demonstrated recently (104). The authors analysed a comprehensive set of environmental variables such as pollution, area deprivation, greenspace and distance to various facilities, and reduced redundancy by applying confirmatory-factor analysis. Thereafter, sparce canonical correlation analysis was employed to identify complex living profiles related to distinct psychiatric symptom groups, while simultaneously allowing the qualitative and quantitative assessment of each factor and their contribution to risk or resilience. Finally, multiple sparse canonical correlation analysis explored the mediating role of brain morphology in the observed associations.

These findings lay the groundwork for understanding the biological processes involved in complex real-life environmental challenges. Further studies are needed to expand upon and provide deeper insides into the specific mechanisms and identify biomarkers for risk and resilience, using deeply phenotyped datasets. Moreover, the applicability of the findings should be examined across diverse populations, settings, and environmental conditions.

### Addressing long latency periods

Long latency periods may exist between exposure to environmental hazards and the onset of mental health or brain outcomes, making it further challenging to establish a clear cause-effect relationship. Most studies are cross-sectional and are based on a single MRI measurement. Repeated measurements across the lifespan could give insights into the temporal relationships and enable the examination of critical periods of vulnerability, windows of intervention, and long-term consequences of early-life exposure on later brain health. Therefore, longitudinal studies are crucial for examining these long-term trajectories of brain development, ageing and degeneration related to environmental exposures. Prominent examples of such studies include IMAGEN (214), ABCD (Adolescent Brain Cognitive Development) (215), Generation R (216), along with the ongoing follow-up assessments in the UK Biobank (217) and the NAKO (German National Cohort) (218). Furthermore, longitudinal studies could help to assess pre- and post-exposure effects on brain outcomes. In this way, the immediate and delayed effects on the brain can be evaluated, as well as the potential reversibility or persistence of these effects.

### Enhancing macroenvironmental exposure assessment

Current literature relies on assessments of environmental factors that are based on a few stations or land use regression models which are spatially and temporally misaligned with the location or period of interest and may not capture accurately the level of environmental exposure. This issue is particularly important when studying susceptibility periods. Environmental exposures often vary in intensity, duration, and timing, posing additional challenges in their accurate measurement. Misclassification of environmental exposures might hinder small but clinically relevant associations or result in spurious associations. To improve the accuracy of exposure assessment, an increased granularity in the spatial and temporal resolution of data collection is required. Remote sensing satellite data, and integration of multiple data sources, such as air quality models and meteorological reanalysis data provide globally standardised environmental measures enabling the tracing of environmental features spanning back several decades (219–221). The wealth of historical environmental data facilitates global comparative analyses and enables the assessment of the cumulative effects of environmental exposures. A recent study among young adults from China and Europe exemplified the application of several satellite-based measures of urbanicity to characterise spatiotemporal patterns of mental disorders risk (121). Confirmatory factor analysis was performed to develop a composite urbanicity measure, which was calculated for each participant from birth to age of recruitment. This approach allowed to assess the cumulative effects and the susceptibility periods of lifetime urban exposure on brain and behaviours.

Measures of urbanicity or other features of macroenvironment that can be applied to different sociocultural conditions and geographies are significant, as they might uncover common associations with brain and behaviour and may assist in global public health policies and urban planning.

### Embracing mobility

Another source of misclassification is the static exposure assessment, disregarding that individuals are exposed to multiple environments along their daily movements. Considering the high spatial and temporal variability of some environmental exposures (e.g., pollutants associated with traffic and industrial production), the actual environmental exposure should be linked to the individual movement patterns and residence time to capture aetiological meaningful associations. Incorporating mobility patterns in data collection, such as daily movements, commuting behaviours, and residential relocations in combination with utilization of geospatial techniques and geographic information systems will allow more accurate assessment of cumulative exposures. Furthermore, leveraging technology, such as wearable devices and mobile applications, alongside ecological momentary assessments, to collect real-time data on individuals’ environmental exposures may be helpful to overcome the ‘static assumption’ errors (222,223). By integrating sensors that measure parameters such as temperature, humidity, UV radiation, air pollution and activity levels, wearable devices provide a personalized perspective into the microclimates individuals experience throughout their daily lives, accounting for factors such as indoor and outdoor environments and personal behaviours. This granular data allows the identification of patterns and correlations between atmospheric variables and their impact on mental well-being (224,225).

### Consolidating future directions

To identify complex real-life environmental profiles and establish their relationship with brain and behaviour, a dataset with adequate overall power is essential. It can be achieved by increasing between-participant variations (combining study populations with heterogeneous macroenvironment and varying mental illness burden) and decreasing random measurement error (utilising objective measures of macroenvironment, repeated measurements and biomarkers). Driven by these objectives, a concerted effort is being made by the environMENTAL consortium, involving multidisciplinary expertise (213). Through the integration of individual cross-sectional and longitudinal cohorts across Europe and beyond, the consortium aims to leverage the strength of existing datasets, which can be enriched with remote sensing, meteorological and air pollution data, and with digital-health tools enabling real-time data collection (i.e., smart phone applications and ecological momentary assessments). Furthermore, combining federated analyses, using the COIN-STAC platform (Collaborative Informatics and Neuroimaging Suite Toolkit for Anonymous Computation) (226) with data harmonisation, and using representational biostatistical models, will enable the identification of impactful environmental signatures that can be evaluated for their replicability and generalisability across study designs, cultural settings, and molecular levels.

## Conclusions

The current review highlights that various macroenvironmental factors, including air pollution, neighbourhood disadvantage, and urbanicity, may alter brain structure and function and, consequently, mental health. Exposure to these factors, particularly during critical periods of development, might have lasting impacts, resulting in heightened risk for a range of mental illnesses. Then again, detrimental effects of urban environment related to higher risk for mental health disorders, like social stress and air pollution, might be attenuated with exposure to natural environments through decreased stress-related activation in brain regions for emotional regulation (135). Similarly, higher safety norms may mitigate the harmful effects of regional socioeconomic adversity on brain and mental health (162).

However, our understanding of these interactions is still evolving and evidence on specific macro-environmental factors, such as climate, noise and light pollution is comparatively sparse. The short-term and long-term effects of the macroenvironment on brain and mental health is elusive and the need for well-designed longitudinal analyses is pressing. The exploration of mediating and moderating factors, that explain these associations, not only in terms of brain but also, lifestyle and social factors, is essential. Additionally, there is a notable lack of studies on subpopulations and vulnerable groups.

By recognizing the impact of environmental factors on brain plasticity processes, policymakers, and healthcare professionals can work towards creating healthier and more supportive environments that promote mental well-being and resilience.

## Data Availability

Details of the search process and included studies can be found in the manuscript.

## Acknowledgements

This work was supported by the European Union–funded Horizon Europe project ‘environMENTAL’ (101057429); and the UK Research and Innovation (UKRI) under the UK government’s Horizon Europe (10041392 and 10038599) to EP, SH, KS and GS; the European Union-funded FP6 Integrated Project ‘IMAGEN’ (Reinforcement-related behaviour in normal brain function and psychopathology) (LSHM-CT-2007-037286); European Research Council Horizon 2020–funded advanced grant ‘STRATIFY’ (695313); the Medical Research Council Grant ‘c-VEDA’ (MR/N000390/1); the the Human Brain Project (HBP SGA 2, 785907, and HBP SGA 3, 945539); the National Institutes of Health (529 R01DA049238); the German Research Foundation (675346); the National Natural Science Foundation of China (82202093); and the Chinese National High-end Foreign Expert Recruitment Plan and the Alexander von Humboldt Foundation to GS; the Shanghai International Science and Technology Partnership Project (21230780200); and the National Natural Science Foundation of China (42175066) to JY.

## Abbreviations

ACC: anterior Cingulate Cortex
ALAN: Artificial Light At Night
CO: Carbon Monoxide
dlPFC: dorsolateral Prefrontal Cortex
DMN: Default Mode Network
FC: Functional Connectivity
GM: Gray Matter
HPA: Hypothalamic-Pituitary-Adrenal
IFG: Inferior Frontal Gyrus
ipRGCs: intrinsically photosensitive Retinal Ganglion Cells
mPFC: medial Prefrontal Cortex
MRI: Magnetic Resonance Imaging
NAc: Nucleus Accumbens
NO2: Nitrogen Dioxide
NOx: Nitrogen Oxides
PAH: Polycyclic Aromatic Hydrocarbons
PFC: Prefrontal Cortex
PM: Particulate Matter
PM10: Particulate Matter with aerodynamic diameter ≤ 10 μm
PM2.5: Particulate Matter with aerodynamic diameter ≤ 2.5 μm
SO2: Sulphur Dioxide
TRAP: Traffic-Related Air Pollution
WH: White Matter

## References

1. Fogarty L, Kandler A. The fundamentals of cultural adaptation: implications for human adaptation. Sci Rep. 2020;10(1):14318.

2. Evans GW. The built environment and mental health. J Urban Health. 2003;80(4):536–55.

3. Lambert KG, Nelson RJ, Jovanovic T, Cerdá M. Brains in the city: Neurobiological effects of urbanization. Neurosci Biobehav Rev. 2015;58:107–22.

4. United Nations, Department of Economic and Social Affairs, Population Division. World Urbanization Prospects: The 2018 Revision [Internet]. New York: United Nations; 2019 [cited 2023 Jun 3]. Available from: https://population.un.org/wup/publications/Files/WUP2018-Report.pdf

5. Ruszkiewicz JA, Tinkov AA, Skalny A V., Siokas V, Dardiotis E, Tsatsakis A, et al. Brain diseases in changing climate. Environ Res. 2019;177:108637.

6. James SL, Abate D, Abate KH, Abay SM, Abbafati C, Abbasi N, et al. Global, regional, and national incidence, prevalence, and years lived with disability for 354 diseases and injuries for 195 countries and territories, 1990&#x2013;2017: a systematic analysis for the Global Burden of Disease Study 2017. Lancet. 2018 Nov 10;392(10159):1789–858.

7. Peen J, Schoevers RA, Beekman AT, Dekker J. The current status of urban-rural differences in psychiatric disorders. Acta Psychiatr Scand. 2010;121(2):84–93.

8. Pedersen CB, Mortensen PB. Evidence of a dose-response relationship between urbanicity during upbringing and schizophrenia risk. Arch Gen Psychiatry. 2001;58(11):1039–46.

9. Attademo L, Bernardini F, Garinella R, Compton MT. Environmental pollution and risk of psychotic disorders: A review of the science to date. Schizophr Res. 2017;181:55–9.

10. Kirkbride J, Coid JW, Morgan C, Fearon P, Dazzan P, Yang M, et al. Translating the epidemiology of psychosis into public mental health: Evidence, challenges and future prospects. J Public Ment Health. 2010;9(2):4–14.

11. Engemann K, Svenning JC, Arge L, Brandt J, Erikstrup C, Geels C, et al. Associations between growing up in natural environments and subsequent psychiatric disorders in Denmark. Environ Res. 2020;188:109788.

12. Hahad O, Lelieveld J, Birklein F, Lieb K, Daiber A, Münzel T. Ambient Air Pollution Increases the Risk of Cerebrovascular and Neuropsychiatric Disorders through Induction of Inflammation and Oxidative Stress. Int J Mol Sci. 2020;21(12):4306.

13. Brook RD, Rajagopalan S, Pope CA, Brook JR, Bhatnagar A, Diez-Roux A V., et al. Particulate Matter Air Pollution and Cardiovascular Disease. Circulation. 2010;121(21):2331–78.

14. Araujo JA. Particulate air pollution, systemic oxidative stress, inflammation, and atherosclerosis. Air Qual Atmos Health. 2011;4(1):79–93.

15. Cipriani G, Danti S, Carlesi C, Borin G. Danger in the Air: Air Pollution and Cognitive Dysfunction. m J Alzheimers Dis Other Demen. 2018;33(6):333–41.

16. da Fonseca ACC, Matias D, Garcia C, Amaral R, Geraldo LH, Freitas C, et al. The impact of microglial activation on blood-brain barrier in brain diseases. Front Cell Neurosci. 2014;8:362.

17. Thomson EM. Air Pollution, Stress, and Allostatic Load: Linking Systemic and Central Nervous System Impacts. Vol. 69, Journal of Alzheimer’s Disease. 2019.

18. Li H, Cai J, Chen R, Zhao Z, Ying Z, Wang L, et al. Particulate matter exposure and stress hormone levels: A randomized, double-blind, crossover trial of air purification. Circulation. 2017;136(7):618–27.

19. Grandjean P, Landrigan PJ. Neurobehavioural effects of developmental toxicity. Lancet Neurol. 2014;13(3):330–8.

20. Binter AC, Kusters MSW, van den Dries MA, Alonso L, Lubczyńska MJ, Hoek G, et al. Air pollution, white matter microstructure, and brain volumes: Periods of susceptibility from pregnancy to preadolescence. Environ Pollut. 2022;313:120109.

21. Guxens M, Lubczyńska MJ, Pérez-Crespo L, Muetzel RL, El Marroun H, Basagaña X, et al. Associations of Air Pollution on the Brain in Children: A Brain Imaging Study. Res Rep Health Eff Inst. 2022;(209):1–61.

22. Guxens M, Lubczyńska MJ, Muetzel RL, Dalmau-Bueno A, Jaddoe VWV, Hoek G, et al. Air Pollution Exposure During Fetal Life, Brain Morphology, and Cognitive Function in School-Age Children. Biol Psychiatry. 2018;84(4):295–303.

23. Lubczyńnska MJ, Muetzel RL, Marroun H El, Basagaña X, Strak M, Denault W, et al. Exposure to air pollution during pregnancy and childhood, and white matter microstructure in preadolescents. Environ Health Perspect. 2020;128(2):27005.

24. Lubczyńska MJ, Muetzel RL, El Marroun H, Hoek G, Kooter IM, Thomson EM, et al. Air pollution exposure during pregnancy and childhood and brain morphology in preadolescents. Environ Res. 2021;198:110446.

25. Peterson BS, Rauh VA, Bansal R, Hao X, Toth Z, Nati G, et al. Effects of prenatal exposure to air pollutants (polycyclic aromatic hydrocarbons) on the development of brain white matter, cognition, and behavior in later childhood. JAMA Psychiatry. 2015;72(6):531–40.

26. Peterson BS, Bansal R, Sawardekar S, Nati C, Elgabalawy ER, Hoepner LA, et al. Prenatal exposure to air pollution is associated with altered brain structure, function, and metabolism in childhood. J Child Psychol Psychiatry. 2022;63(11):1316–31.

27. Beckwith T, Cecil K, Altaye M, Severs R, Wolfe C, Percy Z, et al. Reduced gray matter volume and cortical thickness associated with traffic-related air pollution in a longitudinally studied pediatric cohort. PLoS One. 2020;15(1):e0228092.

28. Mortamais M, Pujol J, Martínez-Vilavella G, Fenoll R, Reynes C, Sabatier R, et al. Effects of prenatal exposure to particulate matter air pollution on corpus callosum and behavioral problems in children. Environ Res. 2019;178:108734.

29. Pujol J, Fenoll R, Macià D, Martínez-Vilavella G, Alvarez-Pedrerol M, Rivas I, et al. Airborne copper exposure in school environments associated with poorer motor performance and altered basal ganglia. Brain Behav. 2016;6(6):e00467.

30. Pujol J, Martínez-Vilavella G, Macià D, Fenoll R, Alvarez-Pedrerol M, Rivas I, et al. Traffic pollution exposure is associated with altered brain connectivity in school children. Neuroimage. 2016;129:175–84.

31. Cserbik D, Chen JC, McConnell R, Berhane K, Sowell ER, Schwartz J, et al. Fine particulate matter exposure during childhood relates to hemispheric-specific differences in brain structure. Environ Int. 2020;143:105933.

32. Miller JG, Dennis EL, Heft-Neal S, Jo B, Gotlib IH. Fine Particulate Air Pollution, Early Life Stress, and Their Interactive Effects on Adolescent Structural Brain Development: A Longitudinal Tensor-Based Morphometry Study. Cereb Cortex. 2022;32(10):2156–69.

33. Alemany S, Vilor-Tejedor N, García-Esteban R, Bustamante M, Dadvand P, Esnaola M, et al. Traffic-Related Air Pollution, APOE ε4 Status, and Neurodevelopmental Outcomes among School Children Enrolled in the BREATHE Project (Catalonia, Spain). Environ Health Perspect. 2019;29(8):087001.

34. Mortamais M, Pujol J, van Drooge BL, Macià D, Martínez-Vilavella G, Reynes C, et al. Effect of exposure to polycyclic aromatic hydrocarbons on basal ganglia and attention-deficit hyperactivity disorder symptoms in primary school children. Environ Int. 2017;105:12–9.

35. Zuo Z, Ran S, Wang Y, Li C, Han Q, Tang Q, et al. Asymmetry in cortical thickness and subcortical volume in treatment-naïve major depressive disorder. Neuroimage Clin. 2019;21:101614.

36. Kong XZ, Postema MC, Guadalupe T, de Kovel C, Boedhoe PSW, Hoogman M, et al. Mapping brain asymmetry in health and disease through the ENIGMA consortium. Hum Brain Mapp. 2022;43(1):167–81.

37. Mundorf A, Peterburs J, Ocklenburg S. Asymmetry in the Central Nervous System: A Clinical Neuroscience Perspective. Front Syst Neurosci. 2021;15:733898.

38. Wilker EH, Preis SR, Beiser AS, Wolf PA, Au R, Kloog I, et al. Long-Term Exposure to Fine Particulate Matter, Residential Proximity to Major Roads and Measures of Brain Structure. Stroke. 2015;46(5):1161–6.

39. Erickson LD, Gale SD, Hedges DW, Brown BL, Anderson JE. Association between exposure to air pollution and total gray matter and total white matter volumes in adults: A cross-sectional study. Brain Sci. 2020;10(3):164.

40. Chen JC, Wang X, Wellenius GA, Serre ML, Driscoll I, Casanova R, et al. Ambient air pollution and neurotoxicity on brain structure: Evidence from women’s health initiative memory study. Ann Neurol. 2015;78(3):466–76.

41. Chen C, Hayden KM, Kaufman JD, Espeland MA, Whitsel EA, Serre ML, et al. Adherence to a MIND-Like Dietary Pattern, Long-Term Exposure to Fine Particulate Matter Air Pollution, and MRI-Based Measures of Brain Volume: The Women’s Health Initiative Memory Study-MRI. Environ Health Perspect. 2021 May 7;129(12):127008.

42. Chen JC, Wang X, Serre M, Cen S, Franklin M, Espeland M. Particulate Air Pollutants, Brain Structure, and Neurocognitive Disorders in Older Women. Res Rep Health Eff Inst. 2017;(193):1–65.

43. Power MC, Lamichhane AP, Liao D, Xu X, Jack CR, Gottesman RF, et al. The association of long-term exposure to particulate matter air pollution with brain MRI findings: The ARIC study. Environ Health Perspect. 2018;126(2):027009.

44. Nußbaum R, Lucht S, Jockwitz C, Moebus S, Engel M, Jöckel KH, et al. Associations of air pollution and noise with local brain structure in a cohort of older adults. Environ Health Perspect. 2020;128(6):67012.

45. Lo CC, Liu WT, Lu YH, Wu D, Wu CD, Chen TC, et al. Air pollution associated with cognitive decline by the mediating effects of sleep cycle disruption and changes in brain structure in adults. Environ Sci Pollut Res Int. 2022;29(35):52355–66.

46. Cho J, Noh Y, Kim SY, Sohn J, Noh J, Kim W, et al. Long-term ambient air pollution exposures and brain imaging markers in Korean adults: The environmental pollution-induced neurological effects (EPINEF) study. Environ Health Perspect. 2020;128(11):117006.

47. Hedges D W., Erickson L D, Gale S D, Anderson J E, Brown B L. Association between exposure to air pollution and thalamus volume in adults: A cross-sectional study. PLoS One. 2020 May;15(3):e0230829.

48. Casanova R, Wang X, Reyes J, Akita Y, Serre ML, Vizuete W, et al. A voxel-based morphometry study reveals local brain structural alterations associated with ambient fine particles in older women. Front Hum Neurosci. 2016;10:495.

49. Gale SD, Erickson LD, Anderson JE, Brown BL, Hedges DW. Association between exposure to air pollution and prefrontal cortical volume in adults: A cross-sectional study from the UK biobank. Environ Res. 2020;185:109365.

50. Petkus AJ, Resnick SM, Wang X, Beavers DP, Espeland MA, Gatz M, et al. Ambient air pollution exposure and increasing depressive symptoms in older women: The mediating role of the prefrontal cortex and insula. Sci Total Environ. 2022;823:153642.

51. Balboni E, Filippini T, Crous-Bou M, Guxens M, Erickson LD, Vinceti M. The association between air pollutants and hippocampal volume from magnetic resonance imaging: A systematic review and meta-analysis. Environ Res. 2022;204(Pt A):111976.

52. Glaubitz L, Stumme J, Lucht S, Moebus S, Schramm S, Jockwitz C, et al. Association between Long-Term Air Pollution, Chronic Traffic Noise, and Resting-State Functional Connectivity in the 1000BRAINS Study. Environ Health Perspect. 2022;130(9):97007.

53. Li Z, Yan H, Zhang X, Shah S, Yang G, Chen Q, et al. Air pollution interacts with genetic risk to influence cortical networks implicated in depression. Proc Natl Acad Sci U S A. 2021;118(46):e2109310118.

54. Mancuso L, Fornito A, Costa T, Ficco L, Liloia D, Manuello J, et al. A meta-analytic approach to mapping co-occurrent grey matter volume increases and decreases in psychiatric disorders. Neuroimage. 2020;222:117220.

55. Piras F, Vecchio D, Kurth F, Piras F, Banaj N, Ciullo V, et al. Corpus callosum morphology in major mental disorders: A magnetic resonance imaging study. Brain Commun. 2021;3(2):fcab100.

56. Price JL, Drevets WC. Neural circuits underlying the pathophysiology of mood disorders. Trends Cogn Sci. 2012;16(1):61–71.

57. Taylor JM, Whalen PJ. Neuroimaging and Anxiety: the Neural Substrates of Pathological and Non-pathological Anxiety. Curr Psychiatry Rep. 2015;17(6):49.

58. Yan CG, Chen X, Li L, Castellanos FX, Bai TJ, Bo QJ, et al. Reduced default mode network functional connectivity in patients with recurrent major depressive disorder. Proc Natl Acad Sci U S A. 2019;116(18):9078–83.

59. Rolls ET. Chapter 1 - The neuroscience of emotional disorders. In: Heilman KM, Nadeau SE, editors. Handbook of Clinical Neurology. Elsevier; 2021. p. 1–26.

60. Goldstein RZ, Volkow ND. Dysfunction of the prefrontal cortex in addiction: Neuroimaging findings and clinical implications. Nat Rev Neurosci. 2011;12(11):652–69.

61. Koob GF, Volkow ND. Neurobiology of addiction: a neurocircuitry analysis. Lancet Psychiatry. 2016;3(8):760–73.

62. Selemon LD, Zecevic N. Schizophrenia: A tale of two critical periods for prefrontal cortical development. Transl Psychiatry. 2015;5(8):e623.

63. Zundel CG, Ryan P, Brokamp C, Heeter A, Huang Y, Strawn JR, et al. Air pollution, depressive and anxiety disorders, and brain effects: A systematic review. Neurotoxicology. 2022;93:272–300.

64. Heo S, Lee W, Bell ML. Suicide and associations with air pollution and ambient temperature: A systematic review and meta-analysis. Int J Environ Res Public Health. 2021;18(14):7699.

65. Braithwaite I, Zhang S, Kirkbride JB, Osborn DPJ, Hayes JF. Air pollution (Particulate matter) exposure and associations with depression, anxiety, bipolar, psychosis and suicide risk: A systematic review and meta-analysis. Environ Health Perspect. 2019;127(12):126002.

66. Antonsen S, Mok PLH, Webb RT, Mortensen PB, McGrath JJ, Agerbo E, et al. Exposure to air pollution during childhood and risk of developing schizophrenia: a national cohort study. Lancet Planet Health. 2020;4(2):e64–73.

67. Arjunan A, Rajan R. Noise and brain. Physiol Behav. 2020;227:113136.

68. Tortorella A, Menculini G, Moretti P, Attademo L, Balducci PM, Bernardini F, et al. New determinants of mental health: the role of noise pollution. A narrative review. International Review of Psychiatry [Internet]. 2022;34(7–8):783–96. Available from: 10.1080/09540261.2022.2095200

69. Slavich GM, Irwin MR. From stress to inflammation and major depressive disorder: A social signal transduction theory of depression. Psychol Bull. 2014;140(3):774–815.

70. Zaman M, Muslim M, Jehangir A. Environmental noise-induced cardiovascular, metabolic and mental health disorders: a brief review. Environ Sci Pollut Res. 2022;29(51).

71. Wang SS, Glied S, Williams S, Will B, Muennig PA. Impact of aeroplane noise on mental and physical health: A quasi-experimental analysis. BMJ Open. 2022;12(5):e057209.

72. Dzhambov AM, Lercher P. Road traffic noise exposure and depression/anxiety: An updated systematic review and meta-analysis. Int J Environ Res Public Health. 2019;16(21):4134.

73. Clark C, Paunovic K. WHO environmental noise guidelines for the European region: A systematic review on environmental noise and quality of life, wellbeing and mental health. Int J Environ Res Public Health. 2018;15(11):2400.

74. Chen Y, Hansell AL, Clark SN, Cai YS. Environmental noise and health in low-middle-income-countries: A systematic review of epidemiological evidence. Environ Pollut. 2023;316(Pt 2):120605.

75. Pérez-Crespo L, Kusters MSW, López-Vicente M, Lubczyńska MJ, Foraster M, White T, et al. Exposure to traffic-related air pollution and noise during pregnancy and childhood, and functional brain connectivity in preadolescents. Environ Int. 2022;164:107275.

76. Simon KR, Merz EC, He X, Noble KG. Environmental noise, brain structure, and language development in children. Brain Lang. 2022;229:105112.

77. Martínez-Vilavella G, Pujol J, Blanco-Hinojo L, Deus J, Rivas I, Persavento C, et al. The effects of exposure to road traffic noise at school on central auditory pathway functional connectivity. Environ Res. 2023;115574.

78. Copenhaver AE, Roberts RC, LeGates TA. Light-dependent effects on mood: Mechanistic insights from animal models. Prog Brain Res. 2022;273(1):71–95.

79. Lyall LM, Wyse CA, Graham N, Ferguson A, Lyall DM, Cullen B, et al. Association of disrupted circadian rhythmicity with mood disorders, subjective wellbeing, and cognitive function: a cross-sectional study of 91 105 participants from the UK Biobank. Lancet Psychiatry. 2018;5(6):507–14.

80. Partonen T, Treutlein J, Alpman A, Frank J, Johansson C, Depner M, et al. Three circadian clock genes Per2, Arntl, and Npas2 contribute to winter depression. Ann Med. 2007;39(3):229–38.

81. Bedrosian TA, Nelson RJ. Influence of the modern light environment on mood. Mol Psychiatry. 2013;18(7):751–7.

82. Alkozei A, Dailey NS, Bajaj S, Vanuk JR, Raikes AC, Killgore WDS. Exposure to Blue Wavelength Light Is Associated With Increases in Bidirectional Amygdala-DLPFC Connectivity at Rest. Front Neurol. 2021;12:625443.

83. Legates TA, Altimus CM, Wang H, Lee HK, Yang S, Zhao H, et al. Aberrant light directly impairs mood and learning through melanopsin-expressing neurons. Nature. 2012;491(7425):594–8.

84. Vandewalle G, Maquet P, Dijk DJ. Light as a modulator of cognitive brain function. Trends Cogn Sci. 2009;13(10):429–38.

85. Min J young, Min K bok. Outdoor light at night and the prevalence of depressive symptoms and suicidal behaviors: A cross-sectional study in a nationally representative sample of Korean adults. J Affect Disord. 2018;227:199–205.

86. Paksarian D, Rudolph KE, Stapp EK, Dunster GP, He J, Mennitt D, et al. Association of Outdoor Artificial Light at Night with Mental Disorders and Sleep Patterns among US Adolescents. JAMA Psychiatry. 2020;77(12):1266–75.

87. Helbich M, Browning MHEM, Huss A. Outdoor light at night, air pollution and depressive symptoms: A cross-sectional study in the Netherlands. Sci Total Environ. 2020;744:140914.

88. Kang SG, Yoon HK, Cho CH, Kwon S, Kang J, Park YM, et al. Decrease in fMRI brain activation during working memory performed after sleeping under 10 lux light. Sci Rep. 2016;6:36731.

89. Halari R, Simic M, Pariante CM, Papadopoulos A, Cleare A, Brammer M, et al. Reduced activation in lateral prefrontal cortex and anterior cingulate during attention and cognitive control functions in medication-naïve adolescents with depression compared to controls. J Child Psychol Psychiatry. 2009;50(3):307–16.

90. Townsend JD, Bookheimer SY, Foland-Ross LC, Moody TD, Eisenberger NI, Fischer JS, et al. Deficits in inferior frontal cortex activation in euthymic bipolar disorder patients during a response inhibition task. Bipolar Disord. 2012;14(4):442–50.

91. Roberts G, Green MJ, Breakspear M, McCormack C, Frankland A, Wright A, et al. Reduced inferior frontal gyrus activation during response inhibition to emotional stimuli in youth at high risk of bipolar disorder. Biol Psychiatry. 2013;74(1):55–61.

92. Stevens AA, Goldman-Rakic PS, Gore JC, Fulbright RK, Wexler BE. Cortical dysfunction in schizophrenia during auditory word and tone working memory demonstrated by functional magnetic resonance imaging. Arch Gen Psychiatry. 1998;55(12):1097–103.

93. Lederbogen F, Haddad L, Meyer-Lindenberg A. Urban social stress – Risk factor for mental disorders. The case of schizophrenia. Environ Pollut. 2013;183:2–6.

94. United Nations, Department of Economic and Social Affairs. World Social Report: Inequality in a rapidly changing world [Internet]. World Social Report. 2020 [cited 2023 Jun 3]. Available from: https://www.un.org/development/desa/dspd/wp-content/uploads/sites/22/2020/02/World-Social-Report2020-FullReport.pdf

95. Sampson L, Ettman CK, Galea S. Urbanization, urbanicity, and depression: A review of the recent global literature. Curr Opin Psychiatry. 2020;33(3):233–44.

96. Peen J, Dekker J, Schoevers RA, ten Have M, Graaf R, Beekman AT. Is the prevalence of psychiatric disorders associated with urbanization? Soc Psychiat Epidemiol. 2007;42(12):984–9.

97. Sui Y, Ettema D, Helbich M. Longitudinal associations between the neighborhood social, natural, and built environment and mental health: A systematic review with meta-analyses. Health Place [Internet]. 2022;77:102893. Available from: https://www.sciencedirect.com/science/article/pii/S135382922200154X

98. Clark C, Myron R, Stansfeld S, Candy B. A systematic review of the evidence on the effect of the built and physical environment on mental health. Vol. 6, Journal of Public Mental Health. 2007.

99. Baxter AJ, Scott KM, Vos T, Whiteford HA. Global prevalence of anxiety disorders: A systematic review and meta-regression. Psychol Med. 2013;43(5):897–910.

100. Van Den Bosch M, Meyer-Lindenberg A. Environmental Exposures and Depression: Biological Mechanisms and Epidemiological Evidence. Annu Rev Public Health. 2019;40:239–59.

101. Leistner C, Menke A. Hypothalamic–pituitary–adrenal axis and stress. Lanzenberger R, Kranz GS, Savic I, editors. Handb Clin Neurol. 2020;175:55–64.

102. Tost H, Champagne FA, Meyer-Lindenberg A. Environmental influence in the brain, human welfare and mental health. Nat Neurosci. 2015;18(10):1421–31.

103. Karatsoreos IN, McEwen BS. Stress and Brain Function. In: Fink G, Pfaff DW, Levine JE, editors. Handbook of Neuroendocrinology. San Diego: Academic Press; 2012. p. 497–507.

104. Xu J, Liu N, Polemiti E, Garcia-Mondragon L, Tang J, Liu X, et al. Effects of urban living environments on mental health in adults. Nat Med. 2023;29(6):1456–67.

105. Kim GW, Jeong GW, Kim TH, Baek HS, Oh SK, Kang HK, et al. Functional neuroanatomy associated with natural and urban scenic views in the human brain: 3.0T functional MR imaging. Korean J Radiol. 2010;11(5):507–13.

106. Bolouki A. Neurobiological effects of urban built and natural environment on mental health: systematic review. Rev Environ Health. 2023;38(1):169–79.

107. Richter A, Al-Bayati M, Paraskevopoulou F, Krämer B, Pruessner JC, Binder EB, et al. Interaction of FKBP5 variant rs3800373 and city living alters the neural stress response in the anterior cingulate cortex. Stress. 2021;24(4):421–9.

108. Lederbogen F, Kirsch P, Haddad L, Streit F, Tost H, Schuch P, et al. City living and urban upbringing affect neural social stress processing in humans. Nature. 2011;474(7352):498–501.

109. Yoshimura S, Okamoto Y, Onoda K, Matsunaga M, Ueda K, Suzuki S ichi, et al. Rostral anterior cingulate cortex activity mediates the relationship between the depressive symptoms and the medial prefrontal cortex activity. J Affect Disord. 2010;122(1–2):76–85.

110. Streit F, Haddad L, Paul T, Frank J, Schäfer A, Nikitopoulos J, et al. A functional variant in the neuropeptide S receptor 1 gene moderates the influence of urban upbringing on stress processing in the amygdala. Stress. 2014;17(4):352–61.

111. Reed JL, D’Ambrosio E, Marenco S, Ursini G, Zheutlin AB, Blasi G, et al. Interaction of childhood urbanicity and variation in dopamine genes alters adult prefrontal function as measured by functional magnetic resonance imaging (fMRI). PLoS One. 2018;13(4):e0195189.

112. Kühn S, Banaschewski T, Bokde ALW, Büchel C, Quinlan EB, Desrivières S, et al. Brain structure and habitat: Do the brains of our children tell us where they have been brought up? Neuroimage. 2020;222:117225.

113. Zhang X, Yan H, Yu H, Zhao X, Shah S, Dong Z, et al. Childhood urbanicity interacts with polygenic risk for depression to affect stress-related medial prefrontal function. Transl Psychiatry. 2021;11(1):522.

114. Haddad L, Schäfer A, Streit F, Lederbogen F, Grimm O, Wüst S, et al. Brain structure correlates of urban upbringing, an environmental risk factor for schizophrenia. Schizophr Bull. 2015;41(1):115–22.

115. Besteher B, Gaser C, Spalthoff R, Nenadić I. Associations between urban upbringing and cortical thickness and gyrification. J Psychiatr Res. 2017;95:114–20.

116. Frissen A, Van Os J, Habets P, Gronenschild E, Marcelis M. No evidence of association between childhood urban environment and cortical thinning in psychotic disorder. PLoS One. 2017;12(1):e0166651.

117. Calem M, Bromis K, McGuire P, Morgan C, Kempton MJ. Meta-analysis of associations between childhood adversity and hippocampus and amygdala volume in non-clinical and general population samples. Neuroimage Clin. 2017;14:471–9.

118. Gorka AX, Hanson JL, Radtke SR, Hariri AR. Reduced hippocampal and medial prefrontal gray matter mediate the association between reported childhood maltreatment and trait anxiety in adulthood and predict sensitivity to future life stress. Biol Mood Anxiety Disord. 2014;4:12.

119. Benedetti F, Radaelli D, Poletti S, Falini A, Cavallaro R, Dallaspezia S, et al. Emotional reactivity in chronic schizophrenia: Structural and functional brain correlates and the influence of adverse childhood experiences. Psychol Med. 2011;41(3):509–19.

120. Thompson PM, Jahanshad N, Ching CRK, Salminen LE, Thomopoulos SI, Bright J, et al. ENIGMA and global neuroscience: A decade of large-scale studies of the brain in health and disease across more than 40 countries. Transl Psychiatry. 2020;10(1):100.

121. Xu J, Liu X, Li Q, Goldblatt R, Qin W, Liu F, et al. Global urbanicity is associated with brain and behaviour in young people. Nat Hum Behav [Internet]. 2022;6(2):279–93. Available from: 10.1038/s41562-021-01204-7

122. Tang IC, Tsai YP, Lin YJ, Chen JH, Hsieh CH, Hung SH, et al. Using functional Magnetic Resonance Imaging (fMRI) to analyze brain region activity when viewing landscapes. Landsc Urban Plan. 2017;162:137–44.

123. Dadvand P, Tischer C, Estarlich M, Llop S, Dalmau-Bueno A, López-Vicente M, et al. Lifelong residential exposure to green space and attention: A population-based prospective study. Environ Health Perspect. 2017;125(9):097016.

124. Saenen ND, Nawrot TS, Hautekiet P, Wang C, Roels HA, Dadvand P, et al. Residential green space improves cognitive performances in primary schoolchildren independent of traffic-related air pollution exposure. Environ Health. 2023;22(1):33.

125. Li H, Ding Y, Zhao B, Xu Y, Wei W. Effects of immersion in a simulated natural environment on stress reduction and emotional arousal: A systematic review and meta-analysis. Front Psychol. 2023;13:1058177.

126. Georgiou M, Morison G, Smith N, Tieges Z, Chastin S. Mechanisms of impact of blue spaces on human health: A systematic literature review and meta-analysis. Int J Environ Res Public Health. 2021;18(5):2486.

127. Egorov AI, Griffin SM, Converse RR, Styles JN, Sams EA, Wilson A, et al. Vegetated land cover near residence is associated with reduced allostatic load and improved biomarkers of neuroendocrine, metabolic and immune functions. Environ Res. 2017;158:508–21.

128. Roe JJ, Ward Thompson C, Aspinall PA, Brewer MJ, Duff EI, Miller D, et al. Green space and stress: Evidence from cortisol measures in deprived urban communities. Int J Environ Res Public Health. 2013;10(9):4086–103.

129. Tang IC, Tsai YP, Lin YJ, Chen JH, Hsieh CH, Hung SH, et al. Using functional Magnetic Resonance Imaging (fMRI) to analyze brain region activity when viewing landscapes. Landsc Urban Plan. 2017;162:137–44.

130. Martínez-Soto J, Gonzales-Santos L, Pasaye E, Barrios FA. Exploration of neural correlates of restorative environment exposure through functional magnetic resonance. Intelligent Buildings International. 2013;5(sup1):10–28.

131. Tani Y, Fujiwara T, Sugihara G, Hanazato M, Suzuki N, Machida M, et al. Neighborhood Beauty and the Brain in Older Japanese Adults. Int J Environ Res Public Health. 2023;20(1):679.

132. Zhang W, He X, Liu S, Li T, Li J, Tang X, et al. Neural correlates of appreciating natural landscape and landscape garden: Evidence from an fMRI study. Brain Behav. 2019;9(7):e01335.

133. Kühn S, Forlim CG, Lender A, Wirtz J, Gallinat J. Brain functional connectivity differs when viewing pictures from natural and built environments using fMRI resting state analysis. Sci Rep. 2021;11(1):4110.

134. Bratman GN, Hamilton JP, Hahn KS, Daily GC, Gross JJ. Nature experience reduces rumination and subgenual prefrontal cortex activation. Proc Natl Acad Sci U S A. 2015;112(28):8567–72.

135. Sudimac S, Sale V, Kühn S. How nature nurtures: Amygdala activity decreases as the result of a one-hour walk in nature. Mol Psychiatry. 2022;27(11):4446–52.

136. Tost H, Reichert M, Braun U, Reinhard I, Peters R, Lautenbach S, et al. Neural correlates of individual differences in affective benefit of real-life urban green space exposure. Nat Neurosci. 2019;22(9):1389–93.

137. Dimitrov-Discher A, Wenzel J, Kabisch N, Hemmerling J, Bunz M, Schöndorf J, et al. Residential green space and air pollution are associated with brain activation in a social-stress paradigm. Sci Rep. 2022;12(1):10614.

138. Dzhambov AM, Bahchevanov KM, Chompalov KA, Atanassova PA. A feasibility study on the association between residential greenness and neurocognitive function in middle-aged Bulgarians. Arh Hig Rada Toksikol. 2019;70(3):173–85.

139. Besser LM, Lovasi GS, Michael YL, Garg P, Hirsch JA, Siscovick D, et al. Associations between neighborhood greenspace and brain imaging measures in non-demented older adults: the Cardiovascular Health Study. Soc Psychiatry Psychiatr Epidemiol. 2021;56(9):1575–85.

140. Crous-Bou M, Gascon M, Gispert JD, Cirach M, Sánchez-Benavides G, Falcon C, et al. Impact of urban environmental exposures on cognitive performance and brain structure of healthy individuals at risk for Alzheimer’s dementia. Environ Int. 2020;138:105546.

141. Phillips JR, Hewedi DH, Eissa AM, Moustafa AA. The Cerebellum and Psychiatric Disorders. Vol. 3, Front Public Health. 2015. p. 66.

142. Van Haren NEM, Schnack HG, Cahn W, Van Den Heuvel MP, Lepage C, Collins L, et al. Changes in cortical thickness during the course of illness in schizophrenia. Arch Gen Psychiatry. 2011;68(9):871–80.

143. Shad MU, Muddasani S, Rao U. Gray matter differences between healthy and depressed adolescents: A voxel-based morphometry study. J Child Adolesc Psychopharmacol. 2012;22(3):190–7.

144. Wylie KP, Tregellas JR. The role of the insula in schizophrenia. Vol. 123, Schizophr Res. 2010. p. 93–104.

145. Kühn S, Düzel S, Eibich P, Krekel C, Wüstemann H, Kolbe J, et al. In search of features that constitute an “enriched environment” in humans: Associations between geographical properties and brain structure. Sci Rep. 2017;7(1):11920.

146. Taniguchi K, Takano M, Tobari Y, Hayano M, Nakajima S, Mimura M, et al. Influence of External Natural Environment Including Sunshine Exposure on Public Mental Health: A Systematic Review. Psychiatry Int [Internet]. 2022;3(1):91–113. Available from: https://www.mdpi.com/2673-5318/3/1/8

147. Gascon M, Mas MT, Martínez D, Dadvand P, Forns J, Plasència A, et al. Mental health benefits of long-term exposure to residential green and blue spaces: A systematic review. Vol. 12, International Journal of Environmental Research and Public Health. 2015.

148. Schneiders J, Drukker M, Van Der Ende J, Verhulst FC, Van Os J, Nicolson NA. Neighbourhood socioeconomic disadvantage and behavioural problems from late childhood into early adolescence. J Epidemiol Community Health. 2003;57(9):699–703.

149. Rudolph KE, Stuart EA, Glass TA, Merikangas KR. Neighborhood disadvantage in context: The influence of Urbanicity on the association between neighborhood disadvantage and adolescent emotional disorders. Soc Psychiatry Psychiatr Epidemiol. 2014;49(3):467–75.

150. Freedman D, Woods GW. Neighborhood Effects, Mental Illness and Criminal Behavior: A Review. J Politics Law. 2013;6(3):1–16.

151. Chang LY, Wang MY, Tsai PS. Neighborhood disadvantage and physical aggression in children and adolescents: A systematic review and meta-analysis of multilevel studies. Aggress Behav. 2016;42(5):441–54.

152. Alderton A, Villanueva K, O’connor M, Boulangé C, Badland H. Reducing inequities in early childhood mental health: How might the neighborhood built environment help close the gap? a systematic search and critical review. Int J Environ Res Public Health. 2019;16(9):1516.

153. Jorgensen NA, Muscatell KA, McCormick EM, Prinstein MJ, Lindquist KA, Telzer EH. Neighborhood disadvantage, race/ethnicity and neural sensitivity to social threat and reward among adolescents. Soc Cogn Affect Neurosci. 2023;18(1):nsac053.

154. Gruebner O, Rapp MA, Adli M, Kluge U, Galea S, Heinz A. Cities and mental health. Dtsch Arztebl Int. 2017;114(8):121–7.

155. Baranyi G, Di Marco MH, Russ TC, Dibben C, Pearce J. The impact of neighbourhood crime on mental health: A systematic review and meta-analysis. Soc Sci Med. 2021;282:114106.

156. Visser K, Bolt G, Finkenauer C, Jonker M, Weinberg D, Stevens GWJM. Neighbourhood deprivation effects on young people’s mental health and well-being: A systematic review of the literature. Soc Sci Med. 2021;270:113542.

157. Finegood ED, Rarick JRD, Blair C. Exploring longitudinal associations between neighborhood disadvantage and cortisol levels in early childhood. Dev Psychopathol. 2017;29(5):1649–62.

158. Hackman D, Betancourt L, Brodsky N, Hurt H, Farah M. Neighborhood disadvantage and adolescent stress reactivity. Front Hum Neurosci. 2012;6:277.

159. Murtha K, Larsen B, Pines A, Parkes L, Moore TM, Adebimpe A, et al. Associations between neighborhood socioeconomic status, parental education, and executive system activation in youth. Cereb Cortex. 2023;33(4):1058–73.

160. Tomlinson RC, Burt SA, Waller R, Jonides J, Miller AL, Gearhardt AN, et al. Neighborhood poverty predicts altered neural and behavioral response inhibition. Neuroimage. 2020;209:116536.

161. Mullins TS, Campbell EM, Hogeveen J. Neighborhood Deprivation Shapes Motivational-Neurocircuit Recruitment in Children. Psychol Sci. 2020;31(7):881–9.

162. Suarez GL, Burt SA, Gard AM, Burton J, Clark DA, Klump KL, et al. The impact of neighborhood disadvantage on amygdala reactivity: Pathways through neighborhood social processes. Dev Cogn Neurosci. 2022;54:101061.

163. Shanmugan S, Wolf DH, Calkins ME, Moore TM, Ruparel K, Hopson RD, et al. Common and dissociable mechanisms of executive system dysfunction across psychiatricdisorders in youth. Am J Psychiatry. 2016;173(5):517–26.

164. Etkin A, Klemenhagen KC, Dudman JT, Rogan MT, Hen R, Kandel ER, et al. Individual differences in trait anxiety predict the response of the basolateral amygdala to unconsciously processed fearful faces. Neuron. 2004;44(6):1043–55.

165. Wolf DH, Satterthwaite TD, Calkins ME, Ruparel K, Elliott MA, Hopson RD, et al. Functional neuroimaging abnormalities in youth with psychosis spectrum symptoms. JAMA Psychiatry. 2015;72(5):456–65.

166. Rakesh D, Seguin C, Zalesky A, Cropley V, Whittle S. Associations Between Neighborhood Disadvantage, Resting-State Functional Connectivity, and Behavior in the Adolescent Brain Cognitive Development Study: The Moderating Role of Positive Family and School Environments. Biol Psychiatry Cogn Neurosci Neuroimaging. 2021;6(9):877–86.

167. Ramphal B, Whalen DJ, Kenley JK, Yu Q, Smyser CD, Rogers CE, et al. Brain connectivity and socioeconomic status at birth and externalizing symptoms at age 2 years. Dev Cogn Neurosci. 2020;45:100811.

168. Saxbe D, Khoddam H, Piero L Del, Stoycos SA, Gimbel SI, Margolin G, et al. Community violence exposure in early adolescence: Longitudinal associations with hippocampal and amygdala volume and resting state connectivity. Dev Sci. 2018;21(6):e12686.

169. Reda MH, Marusak HA, Ely TD, van Rooij SJH, Stenson AF, Stevens JS, et al. Community Violence Exposure is Associated with Hippocampus–Insula Resting State Functional Connectivity in Urban Youth. Neuroscience. 2021;468:149–57.

170. Sripada C, Gard AM, Angstadt M, Taxali A, Greathouse T, McCurry K, et al. Socioeconomic resources are associated with distributed alterations of the brain’s intrinsic functional architecture in youth. Dev Cogn Neurosci. 2022;58:101164.

171. Smith LB, Thelen E. Development as a dynamic system. Trends Cogn Sci. 2003;7(8):343–8.

172. Zugman A, Alliende LM, Medel V, Bethlehem RAI, Seidlitz J, Ringlein G, et al. Country-level gender inequality is associated with structural differences in the brains of women and men. Proc Natl Acad Sci U S A. 2023 May 16;120(20):e2218782120.

173. Sadhu M, Nicholson T de F, Garcia R, Lampley S, Rain M, Fritz A, et al. Relationship between trust in neighbors and regional brain volumes in a population-based study. Psychiatry Res Neuroimaging. 2019;286:11–7.

174. Dumornay NM, Lebois LAM, Ressler KJ, Harnett NG. Racial Disparities in Adversity During Childhood and the False Appearance of Race-Related Differences in Brain Structure. Am J Psychiatry. 2023;180(2):127–38.

175. Gonzalez MZ, Allen JP, Coan JA. Lower neighborhood quality in adolescence predicts higher mesolimbic sensitivity to reward anticipation in adulthood. Dev Cogn Neurosci. 2016;22:48–57.

176. Harnett NG, Wheelock MD, Wood KH, Goodman AM, Mrug S, Elliott MN, et al. Negative life experiences contribute to racial differences in the neural response to threat. Neuroimage. 2019;202:116086.

177. Gonzalez MZ, Beckes L, Chango J, Allen JP, Coan JA. Adolescent neighborhood quality predicts adult dACC response to social exclusion. Soc Cogn Affect Neurosci. 2015;10(7):921–8.

178. Gur RE, Moore TM, Rosen AFG, Barzilay R, Roalf DR, Calkins ME, et al. Burden of Environmental Adversity Associated with Psychopathology, Maturation, and Brain Behavior Parameters in Youths. JAMA Psychiatry. 2019;76(9):966–75.

179. Hunt JFV, Buckingham W, Kim AJ, Oh J, Vogt NM, Jonaitis EM, et al. Association of Neighborhood-Level Disadvantage With Cerebral and Hippocampal Volume [published correction appears in JAMA Neurol. 2020;77(4):527]. JAMA Neurol. 2020;77(4):451–60.

180. Ku BS, Aberizk K, Addington J, Bearden CE, Cadenhead KS, Cannon TD, et al. The Association Between Neighborhood Poverty and Hippocampal Volume Among Individuals at Clinical High-Risk for Psychosis: The Moderating Role of Social Engagement. Schizophr Bull. 2022;48(5):1032–42.

181. Taylor RL, Cooper SR, Jackson JJ, Barch DM. Assessment of Neighborhood Poverty, Cognitive Function, and Prefrontal and Hippocampal Volumes in Children. JAMA Netw Open. 2020;3(11):e2023774.

182. Butler O, Yang XF, Laube C, Kühn S, Immordino-Yang MH. Community violence exposure correlates with smaller gray matter volume and lower IQ in urban adolescents. Hum Brain Mapp. 2018;39(5):2088–97.

183. Hackman DA, Cserbik D, Chen JC, Berhane K, Minaravesh B, McConnell R, et al. Association of Local Variation in Neighborhood Disadvantage in Metropolitan Areas with Youth Neurocognition and Brain Structure. JAMA Pediatr. 2021;175(8):e210426.

184. Krishnadas R, Mclean J, Batty GD, Burns H, Deans KA, Ford I, et al. Socioeconomic deprivation and cortical morphology: Psychological, social, and biological determinants of ill health study. Psychosom Med. 2013;75(7):616–23.

185. Hunt JFV, Vogt NM, Jonaitis EM, Buckingham WR, Koscik RL, Zuelsdorff M, et al. Association of Neighborhood Context, Cognitive Decline, and Cortical Change in an Unimpaired Cohort. Neurology. 2021;96(20):e2500–12.

186. Vargas TG, Damme KSF, Mittal VA. Differentiating distinct and converging neural correlates of types of systemic environmental exposures. Hum Brain Mapp. 2022;43(7):2232–48.

187. Rakesh D, Zalesky A, Whittle S. Assessment of Parent Income and Education, Neighborhood Disadvantage, and Child Brain Structure. JAMA Netw Open. 2022;5(8):e2226208.

188. Rakesh D, Cropley V, Zalesky A, Vijayakumar N, Allen NB, Whittle S. Neighborhood disadvantage and longitudinal brain-predicted-age trajectory during adolescence. Dev Cogn Neurosci. 2021;51:101002.

189. Whittle S, Vijayakumar N, Simmons JG, Dennison M, Schwartz O, Pantelis C, et al. Role of positive parenting in the association between neighborhood social disadvantage and brain development across adolescence. JAMA Psychiatry. 2017;74(8):824–32.

190. Vargas T, Damme KSF, Mittal VA. Neighborhood deprivation, prefrontal morphology and neurocognition in late childhood to early adolescence. Neuroimage. 2020;220:117086.

191. Thompson R, Hornigold R, Page L, Waite T. Associations between high ambient temperatures and heat waves with mental health outcomes: a systematic review. Public Health. 2018 Aug 1;161:171–91.

192. Gao J, Cheng Q, Duan J, Xu Z, Bai L, Zhang Y, et al. Ambient temperature, sunlight duration, and suicide: A systematic review and meta-analysis. Sci Total Environ. 2019;646:1021–9.

193. Liu J, Varghese BM, Hansen A, Xiang J, Zhang Y, Dear K, et al. Is there an association between hot weather and poor mental health outcomes? A systematic review and meta-analysis. Vol. 153, Environment International. 2021.

194. Wahid SS, Raza WA, Mahmud I, Kohrt BA. Climate-related shocks and other stressors associated with depression and anxiety in Bangladesh: a nationally representative panel study. Lancet Planet Health. 2023;7(2):e137–46.

195. Vida S, Durocher M, Ouarda TBMJ, Gosselin P. Relationship between ambient temperature and humidity and visits to Mental Health Emergency Departments in Québec. Psychiatr Serv. 2012;63(11):1150–3.

196. Ding N, Berry HL, Bennett CM. The importance of humidity in the relationship between heat and population mental health: Evidence from Australia. PLoS One. 2016;11(10):e0164190.

197. Lee S, Salvador C, Tuel A, Vicedo-Cabrera AM. Exploring the association between precipitation and hospital admission for mental disorders in Switzerland between 2009 and 2019. PLoS One. 2023 Apr 24;18(4):e0283200.

198. Tapak L, Maryanaji Z, Hamidi O, Abbasi H, Najafi-Vosough R. Investigating the effect of climatic parameters on mental disorder admissions. Int J Biometeorol. 2018;62(12):2109–18.

199. Rotgé JY, Fossati P, Lemogne C. Climate and prevalence of mood disorders: A cross-national correlation study. J Clin Psychiatry. 2014;75(4):408.

200. Estay SA, Ruiz-Aravena M, Baader T, Gotelli M, Heskia C, Olivares JC, et al. Socioeconomic and environmental contexts of suicidal rates in a latitudinal gradient: Understanding interactions to inform public health interventions. J Psychiatr Res. 2022;148:45–51.

201. Zhang R, Volkow ND. Seasonality of brain function: role in psychiatric disorders. Transl Psychiatry. 2023;13(1):65.

202. Jargowsky PA. Disastrous inferences? The ecological fallacy in disaster and emergency management research. In: Disaster and Emergency Management Methods: Social Science Approaches in Application. Routledge; 2021.

203. Song J, Pan R, Yi W, Wei Q, Qin W, Song S, et al. Ambient high temperature exposure and global disease burden during 1990–2019: An analysis of the Global Burden of Disease Study 2019. Sci Total Environ. 2021;787:147540.

204. Zhang X, Chen F, Chen Z. Heatwave and mental health. J Environ Manage. 2023;332:117385.

205. Cruz J, White PCL, Bell A, Coventry PA. Effect of extreme weather events on mental health: A narrative synthesis and meta-analysis for the UK. Int J Environ Res Public Health. 2020;17(22):8581.

206. Charlson F, Ali S, Benmarhnia T, Pearl M, Massazza A, Augustinavicius J, et al. Climate change and mental health: A scoping review. Int J Environ Res Public Health. 2021 May 1;18(9):4486.

207. Cianconi P, Betrò S, Janiri L. The Impact of Climate Change on Mental Health: A Systematic Descriptive Review. Front Psychiatry. 2020;11:74.

208. Batterham PJ, Brown K, Trias A, Poyser C, Kazan D, Calear AL. Systematic review of quantitative studies assessing the relationship between environment and mental health in rural areas. Aust J Rural Health. 2022;30(3):306–20.

209. Jiang Q, Yang X, Liu K, Li B, Li L, Li M, et al. Hyperthermia impaired human visual short-term memory: An fMRI study. Int J Hyperthermia. 2013;29(3):219–24.

210. Sun G, Qian S, Jiang Q, Liu K, Li B, Li M, et al. Hyperthermia-Induced Disruption of Functional Connectivity in the Human Brain Network. PLoS One. 2013;8(4):e61157.

211. Holz NE, Tost H, Meyer-Lindenberg A. Resilience and the brain: a key role for regulatory circuits linked to social stress and support. Mol Psychiatry. 2020;25(2):379–96.

212. Dormann CF, Elith J, Bacher S, Buchmann C, Carl G, Carré G, et al. Collinearity: A review of methods to deal with it and a simulation study evaluating their performance. Ecography. 2013;36(1):27–46.

213. Schumann G, Andreassen OA, Banaschewski T, Calhoun V, Clinton N, Desrivieres S, et al. How can population neuroscience address global environmental challenges to mental health: the environMENTAL-project. JAMA Psychiatry. 2023;(in press).

214. Schumann G, Loth E, Banaschewski T, Barbot A, Barker G, Büchel C, et al. The IMAGEN study: Reinforcement-related behaviour in normal brain function and psychopathology. Mol Psychiatry. 2010;15(12):1128–39.

215. Volkow ND, Koob GF, Croyle RT, Bianchi DW, Gordon JA, Koroshetz WJ, et al. The conception of the ABCD study: From substance use to a broad NIH collaboration. Vol. 32, Developmental Cognitive Neuroscience. 2018.

216. Kooijman MN, Kruithof CJ, van Duijn CM, Duijts L, Franco OH, van IJzendoorn MH, et al. The Generation R Study: design and cohort update 2017. Eur J Epidemiol. 2016;31(12):1243–64.

217. Sudlow C, Gallacher J, Allen N, Beral V, Burton P, Danesh J, et al. UK Biobank: An Open Access Resource for Identifying the Causes of a Wide Range of Complex Diseases of Middle and Old Age. PLoS Med. 2015;12(3).

218. German National Cohort Consortium. The German National Cohort: aims, study design and organization. Eur J Epidemiol. 2014;29(5):371–82.

219. Seltenrich N. Remote-sensing applications for environmental health research [published correction appears in Environ Health Perspect. 2014 Nov;122(11):A296]. Environ Health Perspect. 2014;122(10):A268–75.

220. Hersbach H, Bell B, Berrisford P, Hirahara S, Horányi A, Muñoz-Sabater J, et al. The ERA5 global reanalysis. Q J R Meteorol Soc. 2020 Jul 1;146:1999–2049.

221. Inness A, Ades M, Agustí-Panareda A, Barré J, Benedictow A, Blechschmidt AM, et al. The CAMS reanalysis of atmospheric composition. Atmos Chem Phys. 2019 Mar 20;19:3515–56.

222. Helbich M. Dynamic Urban Environmental Exposures on Depression and Suicide (NEEDS) in the Netherlands: a protocol for a cross-sectional smartphone tracking study and a longitudinal population register study. BMJ Open. 2019 Aug 1;9(8):e030075.

223. Helbich M. Toward dynamic urban environmental exposure assessments in mental health research. Environ Res. 2018;161:129–35.

224. Bernasconi S, Angelucci A, Aliverti A. A Scoping Review on Wearable Devices for Environmental Monitoring and Their Application for Health and Wellness. Vol. 22, Sensors. 2022.

225. Liu S, Schiavon S, Das HP, Jin M, Spanos CJ. Personal thermal comfort models with wearable sensors. Build Environ. 2019;162:106281.

226. Plis SM, Sarwate AD, Wood D, Dieringer C, Landis D, Reed C, et al. COINSTAC: A privacy enabled model and prototype for leveraging and processing decentralized brain imaging data. Front Neurosci. 2016;10:365.

227. Calderón-Garcidueñas L, Mora-Tiscareño A, Ontiveros E, Gómez-Garza G, Barragán-Mejía G, Broadway J, et al. Air pollution, cognitive deficits and brain abnormalities: A pilot study with children and dogs. Brain Cogn. 2008;68(2):117–27.

228. Calderón-Garcidueñas L, Engle R, Antonieta Mora-Tiscareño AM, Styner M, Gómez-Garza G, Zhu H, et al. Exposure to severe urban air pollution influences cognitive outcomes, brain volume and systemic inflammation in clinically healthy children. Brain Cogn. 2011;77(3):345–55.

229. Burnor E, Cserbik D, Cotter DL, Palmer CE, Ahmadi H, Eckel SP, et al. Association of Outdoor Ambient Fine Particulate Matter With Intracellular White Matter Microstructural Properties Among Children. AMA Netw Open. 2021 Dec 9;4(12):e2138300.

230. Cho J, Sohn J, Noh J, Jang H, Kim W, Cho SK, et al. Association between exposure to polycyclic aromatic hydrocarbons and brain cortical thinning: The Environmental Pollution-Induced Neurological EFfects (EPINEF) study. Sci Total Environ. 2020;737:140097.

231. Gawryluk JR, Palombo DJ, Curran J, Parker A, Carlsten C. Brief diesel exhaust exposure acutely impairs functional brain connectivity in humans: a randomized controlled crossover study [published correction appears in Environ Health. 2023 Jan 23;22(1):10]. Environ Health. 2023;22(1):7.

