## Supplementary Information for "How does the macroenvironment influence brain and behaviour – a review of current status and future perspectives"

**Affiliations:**

### Search strategy and study selection

We conducted a literature search on the association between modalities of the macroenvironment and magnetic resonance imaging (MRI)-assessed brain structure and function in PubMed from January 1, 2010, to April 19, 2023. We used predefined search terms with no restrictions applied except for the filter [Humans]. MeSH (Medical Subject Headings) terms and title/abstract text words related to environmental exposures were employed, including urbanisation, air, noise and light pollution, green space, blue space, regional socioeconomic factors, climate, weather extremes, combined with MRI-detected brain changes in structure and function. One reviewer screened the titles and abstracts of the retrieved articles and if a study seemed eligible full texts were reviewed. The reference lists of relevant peer-reviewed systematic reviews (1–15) that were identified in our formal search were additionally hand-searched for relevant literature. Furthermore, studies known to the authors were added. Studies investigating the association between indoor air pollution and occupational hazards with brain plasticity were excluded.

In the following tables (**Supplemental Table 1 – 3**) the full search strategy for PubMed is provided. All references obtained from the search in PubMed were imported in an excel file. An electronic search will first be undertaken with the excel file for identification of irrelevant records by using the terms: adenoma, angiopathy, atherosclero, bacteria, benign, breast, cancer, carcino, cardiac, cerebral palsy, cholera, Crohn, damage, dementia, dengue, diabetes, diarrhea, dysplasia, epilepsy, gut, hyperactivity, infarct, infect, infraction, injury, knee, lobectomy, lung, lymphoma, malaria, malignant, melanoma, metabolic, metastas, mice, microbio, mouse, myocarditis, necrosis, neoplasm, neurodegenerat, oncolog, pain, Parkinson, probiotic, prostate, pulmonary, rodent, sarcomas, sclerosis, stiffness, stimulation, stroke, thrombosis, tinnitus, tumor, tumour, vaccination.

**Supplemental table 1.** Search terms for studies relating to urban and natural environment, air pollution, climate, weather, and regional socioeconomic status.

| | | |
|---|---|---|
| #1 | Environment | “Environment"[Mesh] OR "Geography"[Mesh] OR “Environmental Pollution"[Mesh] OR “physical environment*”[Title/Abstract] OR “living environment*”[Title/Abstract] OR “healthy environment*”[Title/Abstract] OR “unhealthy environment*”[Title/Abstract] OR “environmental factor*”[Title/Abstract] OR “environmental stressor*” [Title/Abstract] OR “environmental risk factor*”[Title/Abstract] OR “environmental adversity”[Title/Abstract] OR “environmental condition*” OR “environmental predictor*”[Title/Abstract] OR “environmental exposure*”[Title/Abstract] OR “environmental pollution”[Title/Abstract] OR “local environment*”[Title/Abstract] OR “environment pollution”[Title/Abstract] |
| #2 | Urbanisation | Urbanization[Mesh] OR "Built Environment"[Mesh] OR "Environment Design"[Mesh] "Urban Population"[Mesh] OR "Suburban Population"[Mesh] OR "Rural Population"[Mesh] OR “built environment”[Title/Abstract] OR "Environment Design"[ Title/Abstract] OR urban*[Title/Abstract] OR city[Title/Abstract] OR cities[Title/Abstract] town*[Title/Abstract] OR rural*[Title/Abstract] OR suburb*[Title/Abstract] OR downtown[Title/Abstract] OR “non-urban”[Title/Abstract] OR “remote communit*”[Title/Abstract] OR “remote area*”[Title/Abstract] OR “remote region*”[Title/Abstract] OR “isolated communit*”[Title/Abstract] OR “isolated area*”[Title/Abstract] OR “isolated region*”[Title/Abstract] OR “gray space*”[Title/Abstract] OR “grey space*”[Title/Abstract] OR “impervious surface*”[Title/Abstract] OR “impervious area*”[Title/Abstract] OR “impervious surface”[Title/Abstract:~2] |
| #3 | Light Pollution | "Light Pollution"[MeSH] OR "Lighting"[Mesh] OR “light pollution”[Title/Abstract] OR “light at night*”[Title/Abstract] OR LAN[Title/Abstract] OR “artificial light*”[Title/Abstract] OR “night light*”[Title/Abstract] OR “light exposure*” [Title/Abstract] OR “outdoor light*”[Title/Abstract] OR “outdoor light”[Title/Abstract:~2] |
| #4 | Noise | ("Noise"[Mesh] OR “noise pollution”[Title/Abstract] OR “acoustic pollution”[Title/Abstract] OR “aeroplane noise”[Title/Abstract:~2] OR “environmental noise*”[Title/Abstract] OR “environment noise*”[Title/Abstract] OR “aircraft noise*”[Title/Abstract] OR “noise* exposure”[Title/Abstract] OR “ambient noise*” |

| | | |
|---|---|---|
| | | [Title/Abstract] OR “transport noise”[Title/Abstract:~2] OR “transportation noise”[Title/Abstract:~2] OR “residential noise”[Title/Abstract:~2] OR “industrial noise”[Title/Abstract:~2] OR “neighborhood noise”[Title/Abstract:~2] OR “neighbourhood noise”[Title/Abstract:~2] OR “noise level*”[Title/Abstract] OR “urban noise”[Title/Abstract:~2] OR “jet noise”[Title/Abstract:~2] OR “noise disturbance*”[Title/Abstract] OR “community noise”[Title/Abstract:~2] OR “vehicle noise”[Title/Abstract:~2] OR “noise metric*”[Title/Abstract] OR “noise exposure*”[Title/Abstract] OR “noise ind*”[Title/Abstract] OR “turbine noise”[Title/Abstract:~2] OR “freeway noise”[Title/Abstract:~2] OR “highway noise”[Title/Abstract:~2] OR “industry noise”[Title/Abstract:~2] OR “road noise”[Title/Abstract:~2] OR “motorway noise”[Title/Abstract:~2]) NOT “occupational”[Title/Abstract] |
| #5 | Green space | "Nature"[Mesh] OR "Forests"[Mesh] OR "Parks, Recreational"[Mesh] OR “natural environment*”[Title/Abstract] OR “nature exposure*”[Title/Abstract] OR “environmental nature”[Title/Abstract:~10] OR “nature-based”[Title/Abstract] OR “natural land*”[Title/Abstract] OR “natural space*”[Title/Abstract] OR “natural area*”[Title/Abstract] OR “natural setting*”[Title/Abstract] OR “nature reserve”[Title/Abstract] OR “green space*”[Title/Abstract] OR greenspace*[Title/Abstract] OR “green area*”[Title/Abstract] OR greenness[Title/Abstract] OR greenery[Title/Abstract] OR vegetation*[Title/Abstract] OR "tree cover"[Title/Abstract:~2] OR "tree canopy"[Title/Abstract:~2] OR "tree density"[Title/Abstract:~2] OR "tree volume"[Title/Abstract:~4] OR "tree area*"[Title/Abstract:] OR "leaf area*"[Title/Abstract] OR "plant area*"[Title/Abstract] OR "park"[Title/Abstract] OR "parks"[Title/Abstract] OR ((forest[Title/Abstract] OR forests[Title/Abstract] OR forest-based[Title/Abstract] OR “forested area*”[Title/Abstract]) NOT (“forest plot*”[Title/Abstract] OR “random forest*”[Title/Abstract])) OR outdoor*[Title/Abstract] OR “open space*”[Title/Abstract] OR “wilderness”[Title/Abstract] OR “wild land”[Title/Abstract] OR “wild space*”[Title/Abstract] OR “woodland*”[Title/Abstract] OR “land use*”[Title/Abstract] OR “land cover”[Title/Abstract] |
| #6 | Blue space | "Water"[Mesh] OR "Rivers"[Mesh] OR "Oceans and Seas"[Mesh] OR "Wetlands"[Mesh] OR “blue space*”[Title/Abstract] OR “bluespace*”[Title/Abstract] OR “blue area*”[Title/Abstract] OR “blue region*”[Title/Abstract] OR “coast*”[Title/Abstract] OR “wetland*”[Title/Abstract] “freshwater*”[Title/Abstract] OR “lake*”[Title/Abstract] OR “blue bodies”[Title/Abstract] OR “water space*”[Title/Abstract] OR “water bodies”[Title/Abstract] OR “water area*”[Title/Abstract] OR “water region*”[Title/Abstract] OR “water feature*”[Title/Abstract] OR “waterway*”[Title/Abstract] OR “water environment*”[Title/Abstract] OR “sea”[Title/Abstract] OR “river”[Title/Abstract] OR “rivers”[Title/Abstract] OR “riverbank*”[Title/Abstract] OR “riverside*”[Title/Abstract] OR “marine area*”[Title/Abstract] OR “marine environment*”[Title/Abstract] OR “marine region*”[Title/Abstract] OR “aquatic area*”[Title/Abstract] OR “aquatic environment*”[Title/Abstract] OR “aquatic region*”[Title/Abstract] OR “waterscapes”[Title/Abstract] OR “inland water”[Title/Abstract:~2] OR “beach*”[Title/Abstract] OR “pond”[Title/Abstract] OR “ponds” [Title/Abstract] |

| | | |
|---|---|---|
| #7 | Regional Socioeconomic status | "Residence Characteristics"[Mesh] OR "Housing/standards"[MAJR] OR "Housing Quality"[Mesh] OR "Social Environment"[Mesh] OR "Neighborhood Characteristics"[Mesh] OR "Poverty Areas"[Mesh] OR "residence characteristic*"[Title/Abstract] OR "residential characteristic*"[Title/Abstract] OR "community characteristic*"[Title/Abstract] OR "community deprivation"[Title/Abstract:~3] OR "residential deprivation"[Title/Abstract:~3] OR "residence deprivation"[Title/Abstract:~3] OR "deprivation index"[Title/Abstract] OR "community deprivation"[Title/Abstract:~2] OR neighborhood*[Title/Abstract] OR neighbourhood*[Title/Abstract] OR "residence area*"[Title/Abstract] OR "residential area*"[Title/Abstract] OR "residential environment"[Title/Abstract] OR "residential deprivation"[Title/Abstract:~2] OR "residence deprivation"[Title/Abstract:~2] OR "area deprivation"[Title/Abstract:~2] OR "deprivation areas"[Title/Abstract:~2] OR "deprivation area"[Title/Abstract:~2] OR ghetto*[Title/Abstract] OR slum*[Title/Abstract] OR "poverty area*"[Title/Abstract] OR "neighborhood poverty"[Title/Abstract:~2] OR "neighbourhood poverty"[Title/Abstract:~2] OR "community poverty"[Title/Abstract:~2] OR "poverty region*"[Title/Abstract] OR "regional poverty"[Title/Abstract:~2] OR "local poverty"[Title/Abstract:~2] OR "poor area*"[Title/Abstract] OR "poor region*"[Title/Abstract] OR "low income area*"[Title/Abstract] OR "low income region*"[Title/Abstract] OR "middle income area*"[Title/Abstract] OR "middle income region*"[Title/Abstract] OR "high income area*"[Title/Abstract] OR "high income region*"[Title/Abstract] OR "social inequ*"[Title/Abstract] OR "economic inequ*"[Title/Abstract] OR "social equit*"[Title/Abstract] OR "economic equit*"[Title/Abstract] OR "social equal*"[Title/Abstract] OR "economic equal*"[Title/Abstract] OR "developing area*"[Title/Abstract] OR "developing region*"[Title/Abstract] OR "local crim*"[Title/Abstract] OR "local crime"[Title/Abstract:~2] OR "area crime"[Title/Abstract:~2] OR "area crim*"[Title/Abstract] OR "community crime"[Title/Abstract:~2] OR "community crim*"[Title/Abstract] OR "crime hot spot*"[Title/Abstract] OR "local safety"[Title/Abstract] OR "area safety"[Title/Abstract:~2] OR "community safety"[Title/Abstract:~2] OR "local violence"[Title/Abstract:~2] OR "area violence"[Title/Abstract:~2] OR "community violence"[Title/Abstract:~2] OR "local socioeconomic"[Title/Abstract:~2] OR "area socioeconomic"[Title/Abstract:~2] OR "community socioeconomic"[Title/Abstract:~2] |
| #8 | Land use/ cover | "Land use"[Title/Abstract] OR "land cover"[Title/Abstract] OR walkability[Title/Abstract] OR walkable[Title/Abstract] |
| #9 | Population density | "Population Density"[Mesh] OR "population densit*"[Title/Abstract] OR overpopulation[Title/Abstract] OR "population size"[Title/Abstract] |
| #10 | Climate/ weather | Climate[Mesh] OR "Climate Change"[Mesh] OR "Extreme Heat"[Mesh] OR "Weather"[Mesh] OR "Extreme Cold Weather"[Mesh] OR "Extreme Hot Weather"[Mesh] OR "Extreme Weather"[Mesh] OR climat*[Title/Abstract] OR weather[Title/Abstract] OR "Extreme Heat"[Title/Abstract] OR "Ambient Temperature*"[Title/Abstract] OR "Air Temperature*"[Title/Abstract] OR "Hot Temperature*"[Title/Abstract] OR "Cold Temperature*"[Title/Abstract] OR "Heat |

| | | |
|---|---|---|
| | | extreme*"[Title/Abstract] OR "Extreme Environment*"[Title/Abstract] OR meteorolog*[Title/Abstract] OR "warming"[Title/Abstract] OR "environmental crisis"[Title/Abstract] OR "climate crisis"[Title/Abstract] OR "greenhouse"[Title/Abstract] OR "heatwave*"[Title/Abstract] OR "heat wave*"[Title/Abstract] OR "cold wave*"[Title/Abstract] |
| #11 | Rainfall | "Humidity"[Mesh] OR "Rain"[Mesh] OR "Snow"[Mesh] OR "rainfall*"[Title/Abstract] OR "rain"[Title/Abstract] OR "humidity"[Title/Abstract] OR "snow"[Title/Abstract] OR "precipitation*"[Title/Abstract] OR "wet day*"[Title/Abstract] OR "Floods"[Mesh] OR "flood*"[Title/Abstract] OR "storm*"[Title/Abstract] |
| #12 | Air pollution | "Air Pollutants"[Mesh] OR "Particulate Matter"[Mesh] OR "Air Pollution"[Mesh] OR "Gases"[Mesh] OR "Volatile Organic Compounds"[Mesh] OR "Sulfur Oxides"[Mesh] OR "Ozone"[Mesh] OR "atmospheric composition"[Title/Abstract] OR "atmosphere composition"[Title/Abstract:~2] OR "air pollutant*"[Title/Abstract] OR "air pollution"[Title/Abstract] OR "particulate matter"[Title/Abstract] OR "PM2.5"[Title/Abstract] OR "PM10"[Title/Abstract] OR "PM(2.5)"[Title/Abstract] OR "PM(10)"[Title/Abstract] OR "ammonia"[Title/Abstract] OR "carbon oxide*"[Title/Abstract] OR "CO2"[Title/Abstract] "CO(2)"[Title/Abstract] OR "carbon monoxide"[Title/Abstract] OR "carbon dioxide"[Title/Abstract] OR "nitrogen oxide*"[Title/Abstract] OR "nitrogen dioxide"[Title/Abstract] OR "NOx"[Title/Abstract] OR "NO2"[Title/Abstract] OR "NO(x)"[Title/Abstract] OR "NO(2)"[Title/Abstract] OR "chlorine"[Title/Abstract] OR "sulfur dioxide"[Title/Abstract] OR "sulphur dioxide"[Title/Abstract] OR "SO2"[Title/Abstract] OR "SO(2)"[Title/Abstract] OR "ozone"[Title/Abstract] OR "O3"[Title/Abstract] OR "volatile organic compound*"[Title/Abstract] OR "VOC"[Title/Abstract] OR "black carbon"[Title/Abstract] OR "polycyclic aromatic hydrocarbons"[Title/Abstract] OR "Aerosol optical depth"[Title/Abstract] OR "AOD"[Title/Abstract] OR "formaldehyde"[Title/Abstract] OR "dust aerosols"[Title/Abstract:~2] OR "particle*"[Title/Abstract] OR "traffic*"[Title/Abstract] OR "ambient*"[Title/Abstract] OR "emission*"[Title/Abstract] OR "outdoor air quality"[Title/Abstract] OR "exhaust gases"[Title/Abstract:~2] OR "vehicle exhaust"[Title/Abstract:~2] OR "exhaust fumes"[Title/Abstract] OR "vehicle fumes"[Title/Abstract:~2] OR "freeway fumes"[Title/Abstract:~2] OR "highway fumes"[Title/Abstract:~2] OR "motorway fumes"[Title/Abstract:~2] OR "road fumes"[Title/Abstract:~2] OR "vehicle gases"[Title/Abstract:~2] OR "freeway gases"[Title/Abstract:~2] OR "highway gases"[Title/Abstract:~2] OR "motorway gases"[Title/Abstract:~2] OR "road gases"[Title/Abstract:~2] |
| #14 | Sunlight/ cloud cover | "Sunlight"[Mesh] OR "sunlight"[Title/Abstract] OR "daylight"[Title/Abstract] OR "sun time"[Title/Abstract:~2] OR "sunshine"[Title/Abstract] OR cloud*[Title/Abstract] OR "sun radiation"[Title/Abstract] |
| #15 | Combine | #1 OR #2 OR #3 OR #4 OR #5 OR #6 OR #7 OR #8 OR #9 OR #10 OR #11 OR #12 OR #13 OR #14 OR #15 |
| #16 | Exclusion criteria | ("Animals"[Mesh] OR "Gastrointestinal Microbiome"[Mesh] OR "Rats"[Mesh] OR COVID-19[Mesh] OR "SARS-CoV-2"[Mesh]) NOT Humans[Mesh] |

| | | |
|---|---|---|
| #17 | Combine | #16 NOT #17 |

**Supplemental table 2.** Search terms for studies relating to MRI-detected changes in brain structure and function.

| | | |
|---|---|---|
| #18 | Brain | (("Brain"[Mesh] OR "Neurosciences"[Mesh] OR “brain”[Title/Abstract] OR “limbic system”[Title/Abstract] OR “neuroscience”[Title/Abstract]) NOT (“brain cancer*”[Title/Abstract] OR “brain tumor*”[Title/Abstract] OR “brain tumour*”[Title/Abstract] OR “brain injur*”[Title/Abstract] OR "Neurodegenerative Diseases"[Mesh] OR "Heredodegenerative Disorders, Nervous System"[Mesh])) |
| #19 | Neuroimaging | "Neuroimaging"[Mesh] OR "Magnetic Resonance Imaging"[Mesh] OR “neuroimag*”[Title/Abstract] OR “brain imag*”[Title/Abstract] OR “diffusion tensor”[Title/Abstract] OR “diffusion tractography”[Title/Abstract] OR “DTI”[Title/Abstract] OR “magnetic resonance”[Title/Abstract] OR “MRI”[Title/Abstract] OR “fMRI*”[Title/Abstract] OR “functional magnetic”[Title/Abstract] OR “sMRI*”[Title/Abstract] OR “structural magnetic”[Title/Abstract] OR “rs-fMRI*”[Title/Abstract] OR “resting state*”[Title/Abstract] OR “FLAIR”[Title/Abstract] OR “3dFLAIR”[Title/Abstract] OR “fluid attenuated inversion recovery”[Title/Abstract] OR “T1-w”[Title/Abstract] OR “T2-w”[Title/Abstract] OR “T1-weight*”[Title/Abstract] OR “T2-weight*”[Title/Abstract] OR “T1W”[Title/Abstract] OR “T2W”[Title/Abstract] OR “weighted imag*”[Title/Abstract] OR “Freesurfer”[Title/Abstract] |
| #20 | Brain areas | “limbic thalam*”[Title/Abstract] OR “Hippocamp*”[Title/Abstract] OR “parahippocamp*”[Title/Abstract] OR “subiculum*”[Title/Abstract] OR ((“CA1”[Title/Abstract] OR “CA-1”[Title/Abstract] OR “CA2”[Title/Abstract] OR “CA-2”[Title/Abstract] OR “CA3”[Title/Abstract] OR “CA-3”[Title/Abstract]) AND (“region*”[Title/Abstract] OR “area*”[Title/Abstract])) OR ((“dentate”[Title/Abstract]) AND (“gyrus”[Title/Abstract] OR “region*”[Title/Abstract] OR “area*”[Title/Abstract] OR “fasc*”[Title/Abstract])) OR “hypothalam*”[Title/Abstract] OR “limbic lobe*”[Title/Abstract] OR “gyrus cinguli”[Title/Abstract] OR “perforant pathway”[Title/Abstract] OR “septum brain”[Title/Abstract:~2] OR “septal nuclei”[Title/Abstract] OR “septum pellucidum”[Title/Abstract] OR “amygdala”[Title/Abstract] OR “basolateral nuclear complex”[Title/Abstract] OR “central amygdaloid nucleus”[Title/Abstract] OR “corticomedial nuclear complex”[Title/Abstract] OR “epithalamus”[Title/Abstract] OR “habenula”[Title/Abstract] OR “pineal gland”[Title/Abstract] OR “insular cortex”[Title/Abstract] OR |

| | | |
|---|---|---|
| | | ((“orbito*”[Title/Abstract] OR “front*”[Title/Abstract] OR “pariet*”[Title/Abstract] OR “tempo*”[Title/Abstract] OR “occipit*”[Title/Abstract] OR “arterio*”[Title/Abstract]) AND (“lobe”[Title/Abstract] OR “region*”[Title/Abstract] OR “area*”[Title/Abstract] OR “cortex”[Title/Abstract] OR “ROI”[Title/Abstract] OR “tissue*”[Title/Abstract])) OR “grey matter”[Title/Abstract] OR “gray matter”[Title/Abstract] OR “white matter”[Title/Abstract] OR “subcortex”[Title/Abstract] OR “sub-cortex”[Title/Abstract] OR “cerebell*”[Title/Abstract] OR “cortical”[Title/Abstract] OR “subcortical”[Title/Abstract] OR “sub-cortical”[Title/Abstract] OR “cerebral”[Title/Abstract] OR “cingulate”[Title/Abstract] OR “gyrus cinguli”[Title/Abstract] OR “neocort*”[Title/Abstract] OR “postrhinal*”[Title/Abstract] OR “perirhinal*”[Title/Abstract] OR “uncinate fasciculus”[Title/Abstract] |
| #21 | Combine | #28 OR #29 OR #30 |

**Supplemental table 3.** Search terms for studies relating to the macroenvironment and MRI-detected changes in brain structure and function.

| | | |
|---|---|---|
| #22 | Environment and brain | #17 AND #21 |
| #23 | Apply filter | #22 AND Humans[Filter] |

### REFERENCES


1. Clifford A, Lang L, Chen R, Anstey KJ, Seaton A. Exposure to air pollution and cognitive functioning across the life course - A systematic literature review. Environ Res. 2016;147:383–98.

2. Hong C, Efferth T. Systematic Review on Post-Traumatic Stress Disorder Among Survivors of the Wenchuan Earthquake. Trauma Violence Abuse. 2016;17(5):542–61.

3. Power MC, Adar SD, Yanosky JD, Weuve J. Exposure to air pollution as a potential contributor to cognitive function, cognitive decline, brain imaging, and dementia: A systematic review of epidemiologic research. Neurotoxicology. 2016;56:235–53.

4. Mothersill O, Donohoe G. Neural effects of social environmental stress – an activation likelihood estimation meta-analysis. Psychol Med. 2016 Jul 24;46(10):2015–23.

5. Misiak B, Stramecki F, Gawęda Ł, Prochwicz K, Sąsiadek MM, Moustafa AA, et al. Interactions Between Variation in Candidate Genes and Environmental Factors in the Etiology of Schizophrenia and Bipolar Disorder: a Systematic Review. Mol Neurobiol. 2018;55(6):5075–100.

6. Buckley L, Broadley M, Cascio CN. Socio-economic status and the developing brain in adolescence: A systematic review. Child Neuropsychol. 2019 Oct 3;25(7):859–84.

7. Shuda Q, Bougoulias ME, Kass R. Effect of nature exposure on perceived and physiologic stress: A systematic review. Complement Ther Med. 2020;53:102514.

8. Lopuszanska U, Samardakiewicz M. The Relationship Between Air Pollution and Cognitive Functions in Children and Adolescents: A Systematic Review. Cogn Behav Neurol. 2020;33(3):157–78.

9. Chandra M, Rai CB, Kumari N, Sandhu VK, Chandra K, Krishna M, et al. Air Pollution and Cognitive Impairment across the Life Course in Humans: A Systematic Review with Specific Focus on Income Level of Study Area. Int J Environ Res Public Health. 2022;19(3):1405.

10. Bolouki A. Neurobiological effects of urban built and natural environment on mental health: systematic review. Rev Environ Health. 2023;38(1):169–79.

11. Balboni E, Filippini T, Crous-Bou M, Guxens M, Erickson LD, Vinceti M. The association between air pollutants and hippocampal volume from magnetic resonance imaging: A systematic review and meta-analysis. Environ Res. 2022;204(Pt A):111976.

12. Sprague NL, Bancalari P, Karim W, Siddiq S. Growing up green: a systematic review of the influence of greenspace on youth development and health outcomes. J Expo Sci Environ Epidemiol. 2022;32(5):660–81.

13. Zundel CG, Ryan P, Brokamp C, Heeter A, Huang Y, Strawn JR, et al. Air pollution, depressive and anxiety disorders, and brain effects: A systematic review. Neurotoxicology. 2022;93:272–300.

14. Fowler CH, Bagdasarov A, Camacho NL, Reuben A, Gaffrey MS. Toxicant exposure and the developing brain: A systematic review of the structural and functional MRI literature. Neurosci Biobehav Rev. 2023;144:105006.

15. de Prado Bert P, Mercader EMH, Pujol J, Sunyer J, Mortamais M. The Effects of Air Pollution on the Brain: a Review of Studies Interfacing Environmental Epidemiology and Neuroimaging. Curr Environ Health Rep. 2018;5(3):351–64.
